# Gastrointestinal Diseases, Genetic Risk, and Incident Dementia: A Prospective Cohort Study in 352,463 Middle-Aged Adults

**DOI:** 10.1101/2022.11.28.22282820

**Authors:** Shuai Yuan, Lintao Dan, Yao Zhang, Jing Wu, Jianhui Zhao, Jie Chen, Xue Li, Miia Kivipelto, Susanna C. Larsson

## Abstract

**Importance:** Although gastrointestinal disease affects gut microbiota and their metabolites associated with dementia risk, the associations between gastrointestinal diseases and the risk of incident dementia are not established.

**Objective:** To examine the associations between 14 gastrointestinal diseases and incident dementia.

**Design, setting, and participants:** Cohort study. This cohort analysis included 352,463 participants free of baseline dementia. Participants joined the UK Biobank study from 2006 to 2010 and were followed up until 2018 or 2021.

**Exposures:** Fourteen baseline gastrointestinal diseases were defined with data from inpatient, primary care, national cancer registries and self-reported data. Polygenic risk score (RPS) and apolipoprotein E (*APOE*) was constructed by individuals’ genetic data.

**Main Outcomes and Measures:** Primary outcome was incident all-cause dementia ascertained via a inpatient, primary care, and national death registries. Secondary outcomes were risk of common subtypes of dementia, including Alzheimer’s disease and vascular dementia as well as early-onset (i.e., with the onset age < 65 years) and late-onset (≥ 65 years) dementia by diagnosis age.

**Results:** During a median follow-up of 12.4 years among 352,463 participants (mean [SD] age, 55.8 [8.1] years; 54.1% were women), 5648 incident dementia cases were diagnosed. Seven gastrointestinal diseases showed significant associations with an increased risk of dementia after controlling covariates and multiple testing. Compared to individuals without gastrointestinal diseases, the risk of dementia increased from 26% (95% confidence interval 13%-41%) for patients with intestinal diverticular disease to 121% (95% confidence interval 57%-211%) for patients with cirrhosis. The associations were different between certain gastrointestinal diseases and dementia by onset age. The associations appeared to be stronger for cirrhosis (*P* = 0.001), irritable bowel syndrome (*P* < 0.001), gastritis and duodenitis (*P* = 0.002), gastroesophageal reflux disease (*P* < 0.001), and peptic ulcer (*P* = 0.030) with early-onset dementia. There were no multiplicative interactions for PRS or *APOE* (*P* > 0.05).

**Conclusions and relevance:** This study found associations of several gastrointestinal diseases with an increased risk of incident dementia, especially early-onset dementia. Our findings imply the great importance of dementia prevention among patients with gastrointestinal diseases.

**Key Points:** *Question:* Are the gastrointestinal diseases associated with risk of incident dementia?

*Findings:* This large-scaled prospective cohort study including 352,463 participants found that seven gastrointestinal diseases were associated with an increased risk of dementia. The associations of cirrhosis, irritable bowel syndrome, gastritis and duodenitis, gastroesophageal reflux disease, and peptic ulcer appeared to be more strongly associated with early-onset dementia compared to late-onset dementia.

*Meaning:* Seven gastrointestinal diseases are associated with an increased risk of incident dementia and appeared to be stronger for early-onset dementia compared to late-onset dementia.

## 1. Background

Due to the population aging, the number of patients with dementia keeps raising and is projected to increase to 152 million by 2050 worldwide.^1^ The increasing prevalence of dementia has generated a tremendous adverse impact on individuals, their families, and the whole society, leading to an economic cost of approximately 1 trillion dollars annually.^1^ Only a few potentially modifiable risk factors have been identified for dementia, such as low education level, traumatic brain injury, smoking, and possibly physical inactivity and air pollution^1^, and the etiological basis of the disease remains largely uncovered.^2^ Thus, it is of urgency to explore other risk factors for dementia with the aims of deepening the understanding of the disease as well as identifying preventive measures.

With increasing data on omics, microbiome in particular, the gut-brain axis hypothesis of ovonic communication between the central nervous system, the enteric nervous system, and gastrointestinal tract has been widely explored.^3^ The development of dementia caused by damage to central nervous cells has been associated with the alteration of gut microbiota^4,5^ and their related metabolites^6-8^. Gut microbiota and gastrointestinal disorders have mutual impacts;^9,10^ thus, gastrointestinal disorders may also influence the risk of dementia.

Population-based observational studies have found an elevated risk of cognitive decline and dementia among patients with inflammatory bowel disease^11^ and liver diseases^12,13^ even though the evidence was inconsistent^14^. Another study observed that treatment with proton pump inhibitors (PPIs), which are widely used for gastrointestinal diseases, was associated with an increased risk of incident dementia.^15^ Given conflicting and limited data as well as unknown associations for other gastrointestinal diseases, we conducted this study to comprehensively investigate the associations of 14 gastrointestinal endpoints with the risk of dementia.

## 2. Methods

### 2.1 Study Population

This study leveraged data from the UK Biobank, which is an ongoing national prospective cohort project that enrolled over 500,000 individuals from in the UK between 2006 and 2010.^16^The UK Biobank study had been approved by the North West–Haydock Research Ethics Committee (REC reference: 21/NW/0157). All participants had signed an electronic consent. In this study, we excluded individuals with baseline dementia or those diagnosed with dementia in the first year of follow-up (*n* = 4852), individuals who developed any studied gastrointestinal diseases after baseline (*n* = 124,454), and individuals without genetic information (*n* = 20,721). In total, 352,463 participants were included in the final analysis (**Supplementary Figure 1**). The study was conducted in accordance with the STROBE checklist.

### 2.2 Gastrointestinal Diseases

The study focused on fourteen gastrointestinal diseases, including gastroesophageal reflux disease, gastritis and duodenitis, celiac disease, Crohn’s disease, ulcerative colitis, intestinal diverticular disease, irritable bowel syndrome, peptic ulcer, pancreatitis (acute and chronic pancreatitis), gallbladder and biliary diseases (cholangitis, cholecystitis, and cholelithiasis), non-alcoholic fatty liver disease, chronic liver cirrhosis, appendicitis, and overall gastrointestinal cancer (esophageal, gastric, small intestinal, colorectal, pancreatic, gallbladder, and hepatic cancers). These gastrointestinal outcomes were ascertained by diagnostic codes from nationwide inpatient dataset, primary care dataset, cancer registries, and self-report diagnosis reviewed by nurses. Detailed diagnostic codes are displayed in **Supplementary Table 1 and 2**. The accuracy of these diagnostic codes has been found to be high (>89%) in the annual report by the Audit Commission for Local Authorities the National Health Service in England and Wales.^17^

### 2.3 Ascertainment of Dementia

Similarly, the incident dementia was ascertained by diagnostic codes from inpatient dataset, primary care dataset, and death registries. Self-reported dementia diagnosis was used to identify and remove individuals with baseline dementia. The diagnostic codes for all-cause dementia had been used in previous studies and shown to be valid (**Supplementary Table 3**).^18^ To differentiate the etiology of dementia subtypes,^19^ two common subtypes, Alzheimer’s disease and vascular dementia, were defined using diagnostic codes shown in **Supplementary Table 3**. We also classified dementia into early-onset (i.e., with the onset age < 65 years) and late-onset (≥ 65 years) dementia by diagnosis age.

### 2.4 Polygenic Risk Score (PRS) and Apolipoprotein E (APOE)

We constructed a PRS for dementia based on 39 well-established genetic variants ^20^ from large-scaled genome-wide association analyses on Alzheimer’s disease ^21-23^. The PRS was generated by multiplying the genotype dosage of each risk allele for each variant by its respective weight and then summing across all variants. The weights given for the score were derived from International Genomics of Alzheimer’s Project (IGAP) studies.^22,24^ Details of used genetic variants are presented in the **Supplementary Table 4**. Of note, *APOE* gene was not included in the PRS.

The *APOE* haplotypes (ε2/ε3/ε4) were genotyped and determined by 2 genetic variants (i.e., rs429358 and rs7412). Participants with 1 or 2 ε4 alleles were defined as *APOE* ε4 carriers and otherwise as *APOE* ε4 noncarriers.

### 2.5 Assessment of Covariates

We collected information on age at recruitment, sex, educational attainment, smoking status, alcohol consumption, physical activity, and family history of dementia from the baseline questionnaires. Body mass index (BMI) were measured by trained nurses at clinical visits.

Diet quality was assessed by a dementia-associated healthy diet score with data from food frequency questionnaires.^25^ The Townsend deprivation index was constructed as a complex indicator of socioeconomic status using the method instructed online (https://biobank.ndph.ox.ac.uk/showcase/label.cgi?id=76). History of hypertension, stroke, and depression were defined by data from the self-reported questionnaires, electronic health-related records, drug prescription, and baseline blood pressure measurement. Regular use of PPIs was recorded in a verbal interview by trained nurses. If covariate information was missing, we applied single imputation method by imputing the median values for continuous variables or applied a most frequently used category for categorical variables (the highest missing rate among covariates was 4.9%). Detailed information and definition of covariates are presented in **Supplementary Table 5**.

### 2.6 Statistical Analysis

The Student’s t-test and chi-square test was used to examine the differences in baseline characteristics by incident disease status for continuous and categorical variables, respectively. Treating age as timescale, the Cox proportional hazard regression was used to estimate the associations of gastrointestinal diseases with the risk of incident dementia and its subtypes. Three models were performed: 1) model 1 adjusted for age and sex; 2) model 2 additionally adjusted for Townsend deprivation index, educational attainment, BMI, physical activity, diet, smoking status, alcohol consumption, baseline hypertension, and baseline stroke; and 3) model 3 further adjusted for PRS. The proportionality of hazards assumption was examined using the Schoenfeld residuals method and found to be satisfied (*P* > 0.11). We examined the associations of gastrointestinal diseases with the risk of dementia subtypes. Heterogeneity between disease subtypes was calculated using the contrast test method.^26^ We calculated the multiplicative interaction and further performed stratification analysis for PRS categories, *APOE* ε4 carrying status, sex, and educational attainment.^25^ To examine the robustness of the results, we performed several sensitivity analyses: 1) the analysis further adjusted for baseline depression; 2) the analysis further adjusted for family history of dementia; 3) the analysis excluding incident cases diagnosed in the first 3 years of follow-up; 4) the analysis including individuals who developed incident gastrointestinal diseases, and 5) the analysis excluding participants with incident Parkinson’s disease, a major neurodegeneration disease. Given PPIs are routinely used among patients with gastrointestinal diseases and has been associated with dementia,^15^ we also performed a sensitivity analysis with further adjustment for PPI use. To rule out the influence of competing risk, we performed a competing risk model to account for the competing risk of death using R package “cmprsk”. We corrected for multiple comparisons with the false discovery rate (FDR) method. All statistical analyses were conducted using R 4.1.2. Two-sided FDR-adjusted *P* value (Q value) < 0.05 were deemed significant.

## 3. Results

During a median follow-up of 12.4 years, 5648 incident dementia cases, including 1453 Alzheimer’s disease and 649 vascular dementia cases, were diagnosed. The baseline characteristics of 352,463 participants by incident dementia are displayed in **Table 1**. Compared with individuals without incident dementia, the group with incident dementia were older, with more males and poorer socioeconomical conditions, and had more unhealthy behaviors and a higher prevalence of baseline gastrointestinal disorders (**Table 1**).

**Table 1.**
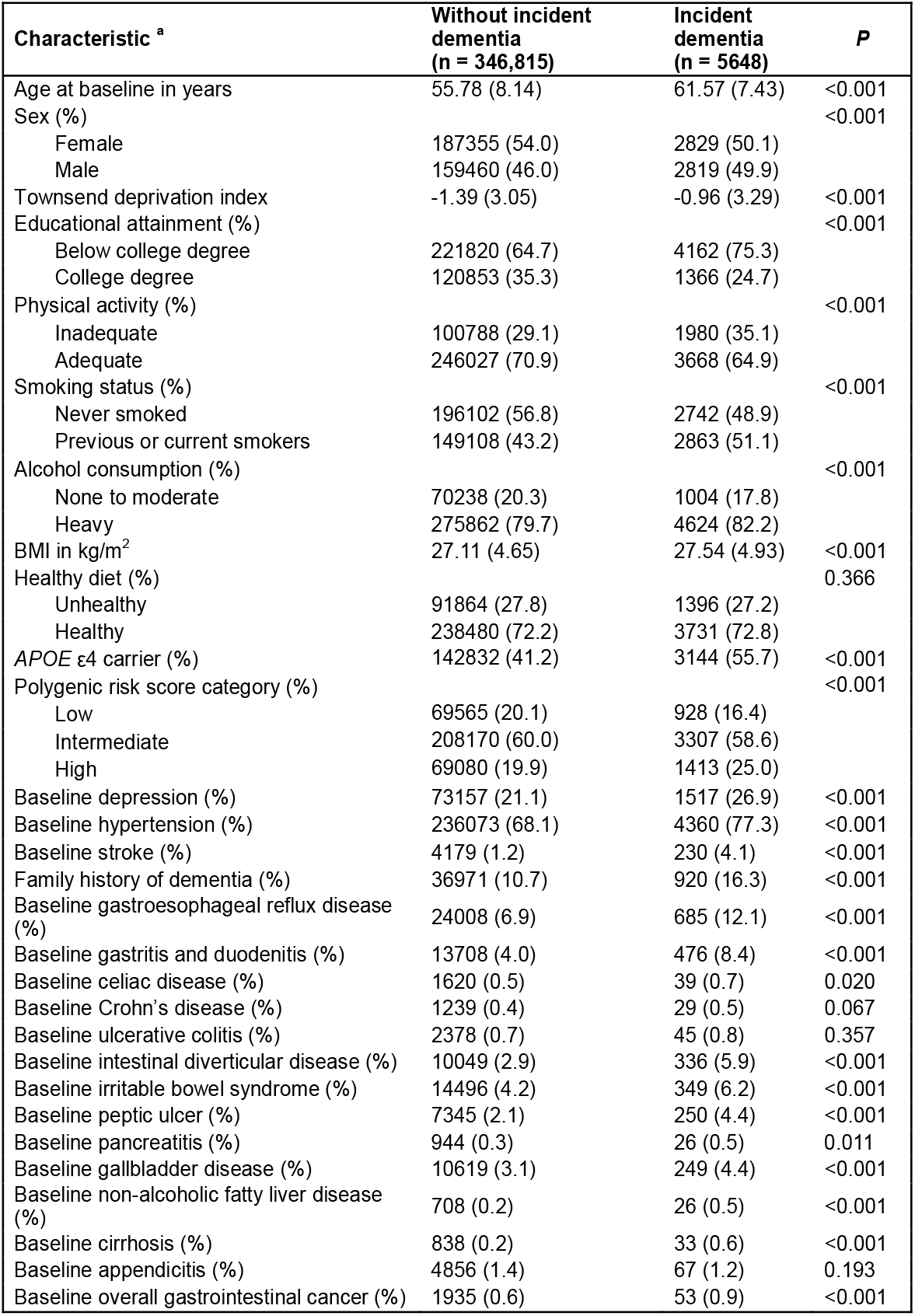
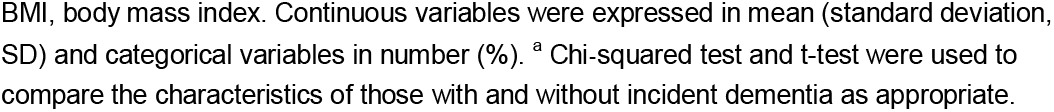
Baseline characteristics by incident dementia.

| **Characteristic <sup>a</sup>** | **Without incident dementia**<br>**(n = 346,815)** | **Incident dementia**<br>**(n = 5648)** | **P** |
| --- | --- | --- | --- |
| Age at baseline in years | 55.78 (8.14) | 61.57 (7.43) | <0.001 |
| Sex (%) |  |  | <0.001 |
| Female | 187355 (54.0) | 2829 (50.1) |  |
| Male | 159460 (46.0) | 2819 (49.9) |  |
| Townsend deprivation index | -1.39 (3.05) | -0.96 (3.29) | <0.001 |
| Educational attainment (%) |  |  | <0.001 |
| Below college degree | 221820 (64.7) | 4162 (75.3) |  |
| College degree | 120853 (35.3) | 1366 (24.7) |  |
| Physical activity (%) |  |  | <0.001 |
| Inadequate | 100788 (29.1) | 1980 (35.1) |  |
| Adequate | 246027 (70.9) | 3668 (64.9) |  |
| Smoking status (%) |  |  | <0.001 |
| Never smoked | 196102 (56.8) | 2742 (48.9) |  |
| Previous or current smokers | 149108 (43.2) | 2863 (51.1) |  |
| Alcohol consumption (%) |  |  | <0.001 |
| None to moderate | 70238 (20.3) | 1004 (17.8) |  |
| Heavy | 275862 (79.7) | 4624 (82.2) |  |
| BMI in kg/m<sup>2</sup> | 27.11 (4.65) | 27.54 (4.93) | <0.001 |
| Healthy diet (%) |  |  | 0.366 |
| Unhealthy | 91864 (27.8) | 1396 (27.2) |  |
| Healthy | 238480 (72.2) | 3731 (72.8) |  |
| APOE ε4 carrier (%) | 142832 (41.2) | 3144 (55.7) | <0.001 |
| Polygenic risk score category (%) |  |  | <0.001 |
| Low | 69565 (20.1) | 928 (16.4) |  |
| Intermediate | 208170 (60.0) | 3307 (58.6) |  |
| High | 69080 (19.9) | 1413 (25.0) |  |
| Baseline depression (%) | 73157 (21.1) | 1517 (26.9) | <0.001 |
| Baseline hypertension (%) | 236073 (68.1) | 4360 (77.3) | <0.001 |
| Baseline stroke (%) | 4179 (1.2) | 230 (4.1) | <0.001 |
| Family history of dementia (%) | 36971 (10.7) | 920 (16.3) | <0.001 |
| Baseline gastroesophageal reflux disease (%) | 24008 (6.9) | 685 (12.1) | <0.001 |
| Baseline gastritis and duodenitis (%) | 13708 (4.0) | 476 (8.4) | <0.001 |
| Baseline celiac disease (%) | 1620 (0.5) | 39 (0.7) | 0.020 |
| Baseline Crohn's disease (%) | 1239 (0.4) | 29 (0.5) | 0.067 |
| Baseline ulcerative colitis (%) | 2378 (0.7) | 45 (0.8) | 0.357 |
| Baseline intestinal diverticular disease (%) | 10049 (2.9) | 336 (5.9) | <0.001 |
| Baseline irritable bowel syndrome (%) | 14496 (4.2) | 349 (6.2) | <0.001 |
| Baseline peptic ulcer (%) | 7345 (2.1) | 250 (4.4) | <0.001 |
| Baseline pancreatitis (%) | 944 (0.3) | 26 (0.5) | 0.011 |
| Baseline gallbladder disease (%) | 10619 (3.1) | 249 (4.4) | <0.001 |
| Baseline non-alcoholic fatty liver disease (%) | 708 (0.2) | 26 (0.5) | <0.001 |
| Baseline cirrhosis (%) | 838 (0.2) | 33 (0.6) | <0.001 |
| Baseline appendicitis (%) | 4856 (1.4) | 67 (1.2) | 0.193 |
| Baseline overall gastrointestinal cancer (%) | 1935 (0.6) | 53 (0.9) | <0.001 |

BMI, body mass index. Continuous variables were expressed in mean (standard deviation, SD) and categorical variables in number (%). <sup>a</sup> Chi-squared test and t-test were used to compare the characteristics of those with and without incident dementia as appropriate.

### 3.1 Gastrointestinal Diseases and Dementia

Eight out of 14 gastrointestinal diseases showed associations with an increased risk of dementia in the model adjusted for age and sex (**Table 2**). After adjustment for covariates and correction for multiple testing, seven associations persisted in model 3 (**Table 2**). In detail, the hazard ratio (HR) of dementia from largest to smallest was 2.21 (95% confidence interval [CI] 1.57, 3.31; *P* < 0.001) for cirrhosis, 2.02 (95% CI 1.38, 2.98; *P* = 0.001) for non-alcoholic fatty liver disease, 1.68 (95% CI 1.53, 1.85; *P* < 0.001) for gastritis and duodenitis, 1.62 (95% CI 1.46, 1.81; *P* < 0.001) for irritable bowel syndrome, 1.48 (95% CI 1.30 1.68; *P* < 0.001) for peptic ulcer, 1.46 (95% CI 1.35, 1.58; *P* < 0.001) for gastroesophageal reflux disease, and 1.26 (95% CI 1.13, 1.41 *P* < 0.001) for intestinal diverticular disease. No association was observed for other gastrointestinal diseases (**Table 2**).

**Table 2.**
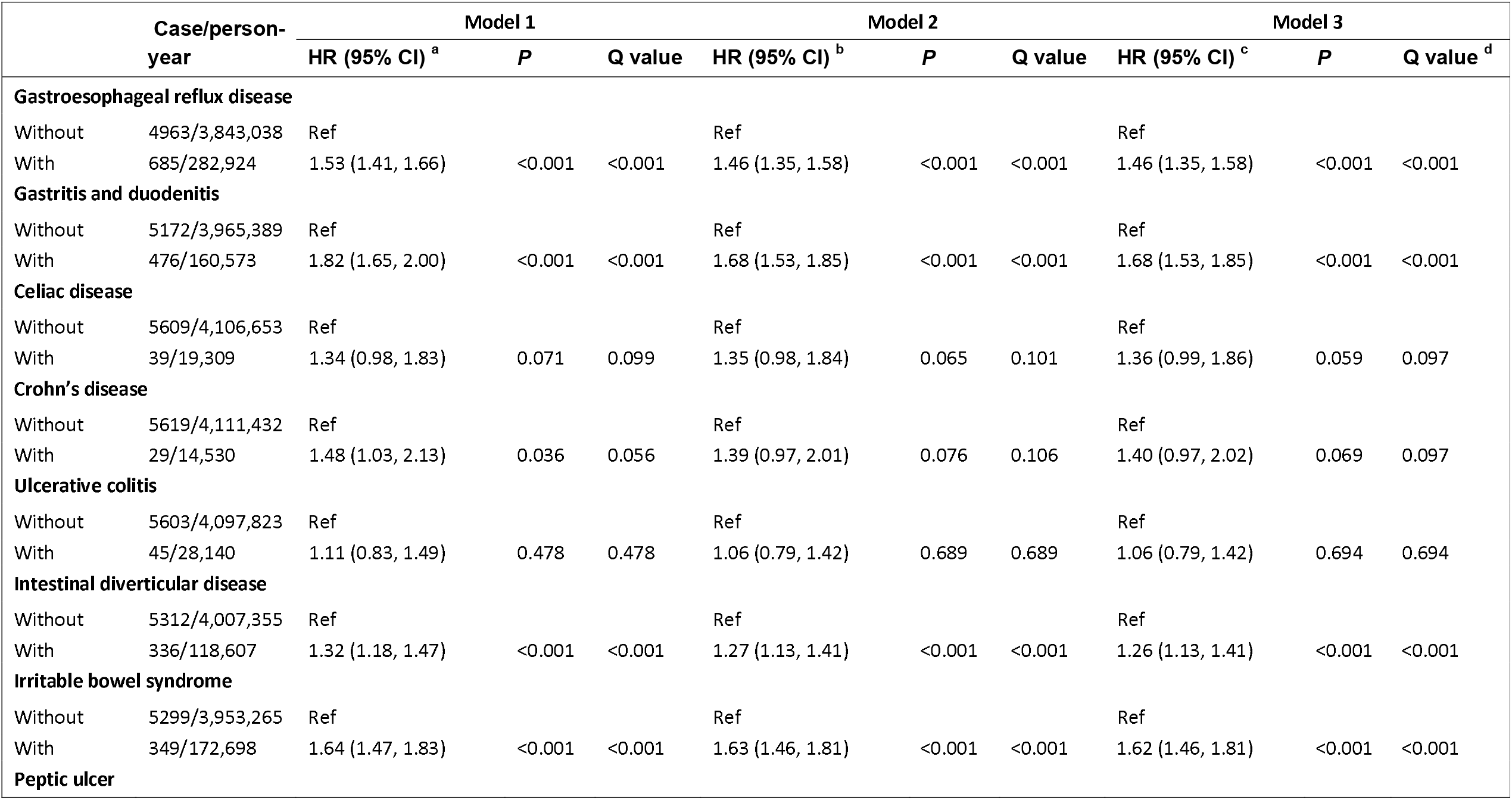
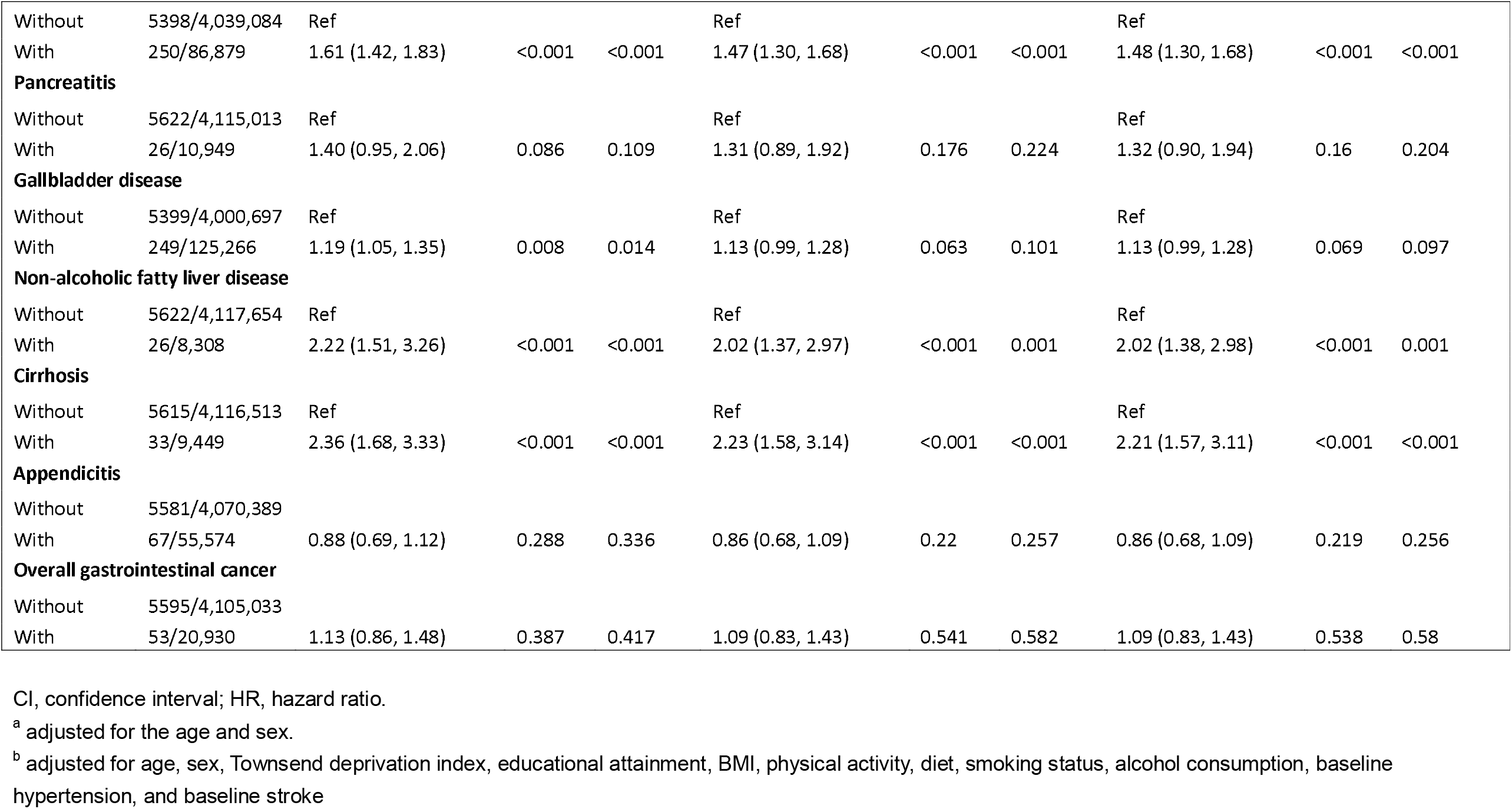
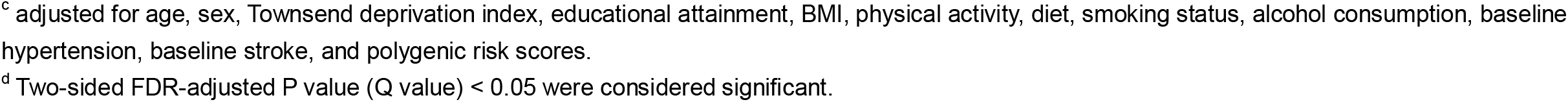
Associations between baseline gastrointestinal diseases and risk of incident dementia.

### 3.2 Gastrointestinal Diseases and Dementia Subtypes

**Table 3** shows the associations of 14 gastrointestinal diseases with risk of incident Alzheimer’s disease and vascular dementia. For Alzheimer’s disease, four associations (irritable bowel syndrome, gastritis and duodenitis, non-alcoholic fatty liver disease, and gastroesophageal reflux disease) were significant in the model 3 after FDR correction. For vascular dementia, five associations (gastritis and duodenitis, peptic ulcer, irritable bowel syndrome, gastroesophageal reflux disease, and intestinal diverticular disease) remained in the model 3 after correction for multiple testing. Gastrointestinal cancer showed a borderline positive association with vascular dementia (HR 1.86; 95% CI 1.07, 3.22; *P* = 0.064), but not with Alzheimer’s disease. No heterogeneity was detected in the associations of gastrointestinal diseases with risk of incident Alzheimer’s disease and vascular dementia (*P*-heterogeneity >0.05).

**Table 3.** Associations between gastrointestinal diseases and risk of incident Alzheimer’s disease and vascular dementia.

|  | Alzheimer's disease |  |  |  | Vascular dementia |  |  |  | *P*-heterogeneity |
| --- | --- | --- | --- | --- | --- | --- | --- | --- | --- |
|  | Case/person-year | HR (95% CI)<sup>a</sup> | *P* | *Q*<sup>b</sup> | Case/person-year | HR (95% CI)<sup>a</sup> | *P* | *Q*<sup>b</sup> |  |
| **Gastroesophageal reflux disease** |  |  |  |  |  |  |  |  | 0.226 |
| Without | 1282/3,857,645 | Ref |  |  | 557/3,857,645 | Ref |  |  |  |
| With | 171/285,279 | 1.30 (1.11, 1.53) | 0.001 | 0.009 | 92/285,279 | 1.54 (1.23, 1.92) | <0.001 | 0.001 |  |
| **Gastritis and duodenitis** |  |  |  |  |  |  |  |  | 0.454 |
| Without | 1337/3,980,771 | Ref |  |  | 585/3,980,771 | Ref |  |  |  |
| With | 116/162,153 | 1.47 (1.22, 1.78) | <0.001 | 0.001 | 64/162,153 | 1.66 (1.28, 2.15) | <0.001 | 0.001 |  |
| **Celiac disease** |  |  |  |  |  |  |  |  | 0.106 |
| Without | 1440/4,123,458 | Ref |  |  | 648/4,123,458 | Ref |  |  |  |
| With | 13/19,466 | 1.64 (0.95, 2.84) | 0.074 | 0.130 | 1/19,466 | 0.30 (0.04, 2.13) | 0.228 | 0.400 |  |
| **Crohn's disease** |  |  |  |  |  |  |  |  | NA |
| Without | 1445/4,128,332 | Ref |  |  | 649/4,128,332 | Ref |  |  |  |
| With | 8/14,592 | 1.54 (0.77, 3.08) | 0.224 | 0.314 | 0/14,592 | NA | NA | NA |  |
| **Ulcerative colitis** |  |  |  |  |  |  |  |  | 0.898 |
| Without | 1444/4,114,682 | Ref |  |  | 645/4,114,682 | Ref |  |  |  |
| With | 9/28,242 | 0.81 (0.42, 1.57) | 0.540 | 0.587 | 4/28,242 | 0.75 (0.28, 2.01) | 0.570 | 0.726 |  |
| **Intestinal diverticular disease** |  |  |  |  |  |  |  |  | 0.277 |
| Without | 1355/4,023,486 | Ref |  |  | 594/4,023,486 | Ref |  |  |  |
| With | 98/119,438 | 1.23 (1.00, 1.51) | 0.050 | 0.119 | 55/119,438 | 1.49 (1.13, 1.97) | 0.005 | 0.014 |  |
| **Irritable bowel syndrome** |  |  |  |  |  |  |  |  | 0.301 |
| Without | 1377/3,968,630 | Ref |  |  | 611/3,968,630 | Ref |  |  |  |
| With | 76/174,294 | 1.39 (1.10, 1.75) | 0.006 | 0.026 | 38/174,294 | 1.72 (1.23, 2.39) | 0.001 | 0.005 |  |
| **Peptic ulcer** |  |  |  |  |  |  |  |  | 0.125 |
| Without | 1392/4,055,330 | Ref |  |  | 606/4,055,330 | Ref |  |  |  |
| With | 61/87,594 | 1.29 (1.00, 1.67) | 0.051 | 0.119 | 43/87,594 | 1.77 (1.30, 2.42) | <0.001 | 0.002 |  |
| **Pancreatitis** |  |  |  |  |  |  |  |  | 0.523 |
| Without | 1443/4,131,905 | Ref |  |  | 646/4,131,905 | Ref |  |  |  |
| With | 10/11,019 | 1.80 (0.97, 3.36) | 0.064 | 0.127 | 3/11,019 | 1.18 (0.38, 3.67) | 0.775 | 0.898 |  |
| **Gallbladder disease** |  |  |  |  |  |  |  |  | 0.160 |
| Without | 1397/4,016,869 | Ref |  |  | 616/4,016,869 | Ref |  |  |  |
| With | 56/126,055 | 0.88 (0.67, 1.15) | 0.351 | 0.447 | 33/126,055 | 1.21 (0.85, 1.72) | 0.288 | 0.403 |  |
| **Non-alcoholic fatty liver disease** |  |  |  |  |  |  |  |  | 0.726 |
| Without | 1445/4,134,532 | Ref |  |  | 646/4,134,532 | Ref |  |  |  |
| With | 8/8,392 | 2.41 (1.20, 4.82) | 0.013 | 0.047 | 3/8,392 | 1.90 (0.61, 5.91) | 0.268 | 0.403 |  |
| **Cirrhosis** |  |  |  |  |  |  |  |  | 0.589 |
| Without | 1446/4,133,368 | Ref |  |  | 647/4,133,368 | Ref |  |  |  |
| With | 7/9,556 | 1.79 (0.85, 3.77) | 0.124 | 0.192 | 2/9,556 | 1.16 (0.29, 4.65) | 0.834 | 0.898 |  |
| **Appendicitis** |  |  |  |  |  |  |  |  | 0.110 |
| Without | 1433/4,087,141 | Ref |  |  | 645/4,087,141 | Ref |  |  |  |
| With | 20/55,783 | 1.07 (0.69, 1.66) | 0.775 | 0.775 | 4/55,783 | 0.45 (0.17, 1.19) | 0.107 | 0.214 |  |
| **Overall gastrointestinal cancer** |  |  |  |  |  |  |  |  | 0.053 |
| Without | 1441/4,121,906 | Ref |  |  | 636/4,121,906 | Ref |  |  |  |
| With | 12/21,018 | 0.84 (0.48, 1.48) | 0.545 | 0.587 | 13/21,018 | 1.86 (1.07, 3.22) | 0.027 | 0.064 |  |

CI, confidence interval; HR, hazard ratio; NA, not applicable.
<sup>a</sup> Associations were derived from model 3 adjusted for age, sex, Townsend deprivation index, educational attainment, BMI, physical activity, diet, smoking status, alcohol consumption, baseline hypertension, and baseline stroke, and polygenic risk score.
<sup>b</sup> Two-sided FDR-adjusted P value (Q value) < 0.05 were considered significant.

The associations of gastrointestinal diseases differed with between early- and late-onset dementia (**Figure 1**). Compared to that for late-onset dementia, the associations for early-onset dementia appeared to be stronger with cirrhosis (HR 4.64; 95% CI 2.83, 7.60; *P* < 0.001), irritable bowel syndrome (HR 2.47; 95% CI 2.09, 2.93; *P* < 0.001), gastritis and duodenitis (HR 2.33; 95% CI 1.92, 2.82; *P* < 0.001), gastroesophageal reflux disease (HR 2.23; 95% CI 1.91, 2.61; *P* < 0.001), and peptic ulcer (HR 1.95; 95% CI 1.48, 2.58; *P* < 0.001). Heterogeneity was detected for these associations between early- and late-onset dementia (*P*-heterogeneity < 0.05).

**Figure 1.**
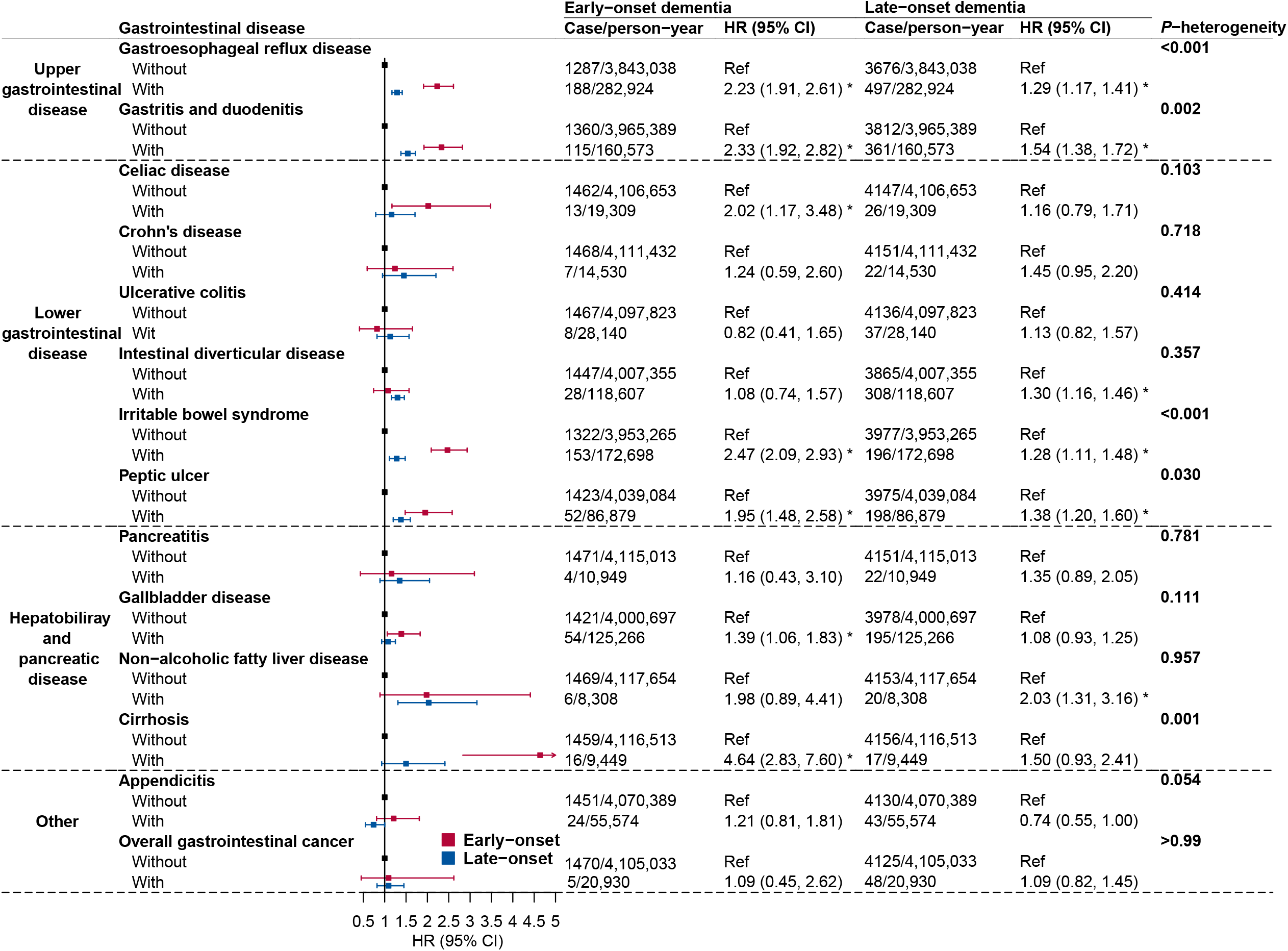
Associations between baseline gastrointestinal diseases and risk of incident dementia by onset age. CI, confidence interval; HR, hazard ratio. Estimates were adjusted for age, sex, Townsend deprivation index, educational attainment, BMI, physical activity, diet, smoking status, alcohol consumption, baseline hypertension, baseline stroke, and polygenic risk score. * Representing a significant association after FDR correction for multiple comparison. *P* for heterogeneity was two-sided FDR-adjusted.

### 3.3 Interactions with Sex, Educational Attainment, PRS, and APOE ε4

There were no interactions between gastrointestinal diseases and sex or between gastrointestinal diseases and educational attainment (*P-interaction* > 0.05, **Supplementary Table 6**). PRS categories were significantly associated with the risk of dementia (**Supplementary Table 7**). We detected no interactions between gastrointestinal diseases and PRS or gastrointestinal diseases and *APOE* ε*4 carrying* status (*P-interaction* > 0.05, **Figure S2 and Figure S3**).

### 3.4 Sensitivity Analyses

The associations remained after accounting for competing risk of death (**Supplementary Table 8**), in a series of other sensitivity analyses (**Supplementary Table 9**), in the analysis including individuals who developed subsequent gastrointestinal diseases after baseline (**Supplementary Table 10**), and in the analysis excluding individuals with incident Parkinson’s disease (**Supplementary Table 11**). Certain associations attenuated slightly albeit remained significant in the sensitivity analysis with further adjustment for regular PPIs use (**Supplementary Table 9**).

## 4. Discussion

This large-scaled prospective cohort study found that seven gastrointestinal diseases were associated with an increased risk of dementia. Compared to individuals without gastrointestinal disorders, the risk of dementia increased from 26% for patients with intestinal diverticular disease to 121% for patients with cirrhosis. As for dementia subtypes, the associations were similar for Alzheimer’s disease and vascular dementia. However, the associations of cirrhosis, irritable bowel syndrome, gastritis and duodenitis, gastroesophageal reflux disease, and peptic ulcer appeared to be more strongly associated with early-onset dementia compared to late-onset dementia. There were no interactions of gastrointestinal diseases with sex, education attainment, genetic risk, or *APOE* ε*4*.

The associations between different gastrointestinal disorders and dementia have been investigated in some previous studies mainly focusing on liver disease,^12-14^ irritable bowel syndrome,^27^ gastritis,^28^ gastroesophageal reflux disease,^29^ and diverticular disease^30^. Most these studies found positive links between gastrointestinal diseases and the risk of dementia, which is in line with our findings based on a large-scaled cohort including more than 6000 incident cases. However, most of the above studies were based on the East Asian population^11,27,30^ that owns a different patten of gastrointestinal diseases compared to other populations due to dietary habits and a high prevalence of *Helicobacter pylori* infection. Our data thus added novel information in support of the positive associations between a broad range of gastrointestinal diseases and dementia risk in the western world. Another study found that inflammatory bowel disease was associated with an increased risk of dementia, in particular early-onset type.^11^ In our study, a borderline positive association was observed for Crohn’s disease. Considering the wide confidence interval of the association, this nonsignificant result might be caused by an inadequate power due to a small number of cases. In addition, the associations of non-alcoholic fatty liver disease and cirrhosis with dementia were inconsistent between studies.^12-14^ Even though our study detected the positive associations between two liver diseases and dementia, a Mendelian randomization analysis revealed a potential inverse effect of non-alcoholic fatty liver disease on dementia in a Finnish cohort.^31^ The reason for this discrepancy is unclear but possibly related to competing risk of death. There are limited studies that examined the associations of gastritis and duodenitis, peptic ulcer, and gallbladder disease with dementia risk. Thus, our novel findings need further verification.

We examined the differences in the associations between gastrointestinal diseases with two common subtypes of dementia. No large differences were observed between the associations of Alzheimer’s disease and vascular dementia with gastrointestinal diseases except for gastrointestinal cancer. In detail, baseline gastrointestinal cancer is associated with an increased risk of vascular dementia but not with Alzheimer’s disease. Even though previous studies found an inverse association of overall cancer diagnosis with the risk of dementia, which might be immune to diagnostic bias, competing risk bias, and confounding.^32^ A higher risk of dementia was observed among colorectal cancer survivors^33^. Another interesting finding of our study in the stronger associations of certain gastrointestinal diseases with early-onset dementia compared to late-onset dementia. The possible reason for the differences may be that early-onset dementia is mainly caused by vascular comorbidities^34^ that have a high prevalence rate among patients with hepatic,^35^ intestinal,^36^ and esophageal^37^ dysfunction. These findings not only deepen the understanding in the etiological differences between early- and late-onset dementia but also may guide the secondary prevention for patients with different gastrointestinal conditions. We did not detect any interaction effects of sex, education attainment, genetic risk, or *APOE* ε*4*, which suggested that the associations between gastrointestinal disease and dementia risk might not differ between sub-populations stratified by these factors. Thus, the corresponding dementia prevention should be promoted in overall population.

The underlying pathways linking gastrointestinal diseases to dementia remain to be established; however, there are several plausible mechanisms that may explain the observed positive associations. The most important bridge of the associations is gut microbiome. The onset of gastrointestinal diseases that alters the normal function and microenvironment of the gastrointestinal tract may impact the diversity, components, and intensity of gut microbiota and the levels of their metabolites.^38^ Animal studies have found that fecal microbiota transplantation between Alzheimer’s disease mouse and healthy controls changed the levels of amyloid and tau, memory function and neurogenesis.^4,39^ In addition, the metabolites of gut microbiota, like bile acid and indole-3 propionate, have been identified as a potential mediator in the associations with cognitive impairment and nerve repair.^6,7,40^ Treatments of gastrointestinal diseases, such as PPIs positively associated with dementia,^15^ may also explain a part of the associations, which is partly supported by our data where most associations slightly attenuated with further adjustment for regular PPI use. *Helicobacter pylori* infection that was a frequent pathogen of gastric disease may also play a role;^28^ however, this appears not to be a strong pathway since the association between *Helicobacter pylori* infection and dementia risk is undetermined^41,42^ and the prevalence of *Helicobacter pylori* infection is quite low in the U.K. For vascular morbidity related gastrointestinal diseases, the increased burden of stroke may mediate the associations with in particular early-onset and vascular dementia. For gastritis and duodenitis and irritable bowel syndrome that have inflammation as a key pathogenic basis, cumulative chronic inflammation may increase the risk of dementia by facilitating neurocognitive changes and subsequent functional decline.^43^ The strengths of the study include a large sample size with a long follow-up time, a comprehensive investigation on a wide range of gastrointestinal conditions, and careful consideration and adjustment for confounders. Nonetheless, several limitations should be noted when interpreting our data. First, this is an observational cohort study, which cannot infer causality. However, we minimized possible biases from reverse causation and residual confounding by excluding incident cases diagnosed during the first year of follow-up and adjusted for vital risk factors for dementia. Second, there might be misclassifications of gastrointestinal diseases even though the used diagnostic codes have been found to be valid ^17^. Due to the prospective design of the study, these misclassifications should be nondifferential and therefore attenuated the associations in a conservative way. Third, except for PPIs, we did not take other treatments for gastrointestinal disorders into consideration. Forth, we might have had insufficient power to detect weak associations owing to few cases for infrequent gastrointestinal diseases. Fifth, a genome-wide cross-trait analysis revealed shared genetic architecture between Alzheimer’s disease and gastrointestinal diseases.^44^ Thus, we could not completely rule out the possibility that the observed associations were caused by shared genetic risk factors even though our results were robust in the analysis with the additional adjustment for the family history of dementia and PRS.

In summary, this cohort study found associations of seven gastrointestinal diseases with an increased risk of incident dementia. The associations appeared to be stronger for early-onset dementia compared to late-onset dementia. Our findings provide novel understandings about the etiology of early- and late-onset dementia and imply the great importance of dementia prevention among patients with gastrointestinal diseases.

## Data Availability

The datasets analysed during the current study are available in a public, open access repository (https://www.ukbiobank.ac.uk/).

## Acknowledgments

We are much obliged to the administer team of the UK Biobank as well as all the participants.

## Ethics approval and consent to participate

The ethical approval was granted for the UK Biobank by the North West-Haydock Research Ethics Committee (REC reference: 21/NW/0157). All participants provided informed consent through electronic signature at baseline assessment. This study was conducted with the UK Biobank Resource under application number 66354.

## CRediT authorship contribution statement

All authors read and approved the final manuscript.

Shuai Yuan (Conceptualization: Supporting; Methodology: Equal; Writing - review & editing: Leading)

Lintao Dan (Conceptualization: Supporting; Methodology: Equal; Formal analysis: Leading; Writing - original draft: Supporting; and Writing - review & editing: Equal)

Yao Zhang (Conceptualization: Supporting; Writing - original draft: Equal; Writing - review & editing: Equal)

Jing Wu (Conceptualization: Supporting; Methodology: Supporting; Writing - review & editing: Equal)

Jianhui Zhao (Conceptualization: Supporting; Writing - review & editing: Equal)

Jie Chen (Conceptualization: Leading; Methodology: Equal; Formal analysis: Supporting; Writing - original draft: Supporting; Writing - review & editing: Equal)

Xue Li (Conceptualization: Leading; Data curation: Equal; and Funding acquisition: Leading; Writing - review & editing: Equal)

Miia Kivipelto (Conceptualization: Supporting; Writing - review & editing: Equal)

Susanna C. Larsson (Conceptualization: Equal; Data curation: Equal; and Funding acquisition: Equal; and Writing - review & editing: Leading)

---

**STROBE Statement—Checklist of items that should be included in reports of *cohort studies***

*Give information separately for exposed and unexposed groups.

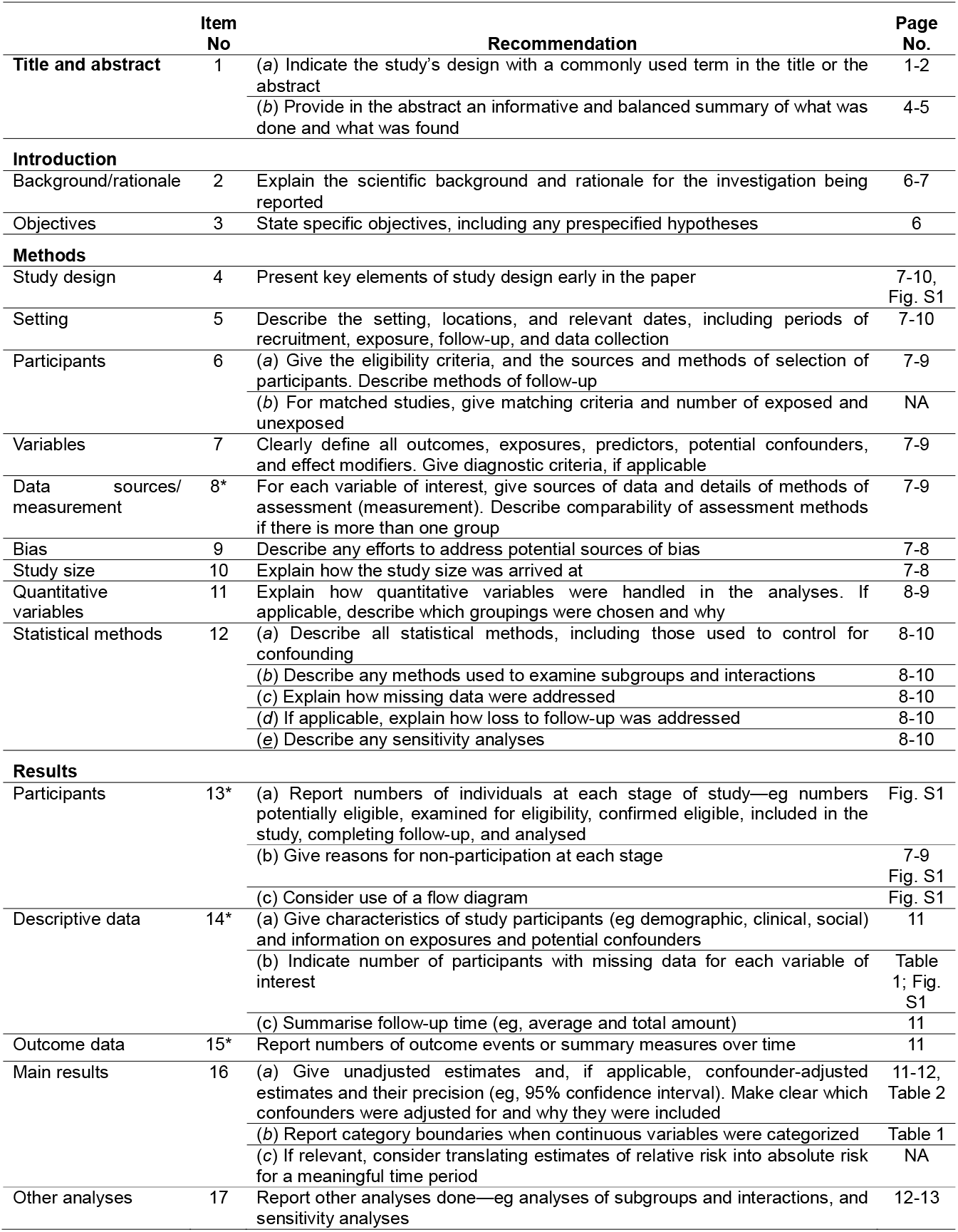
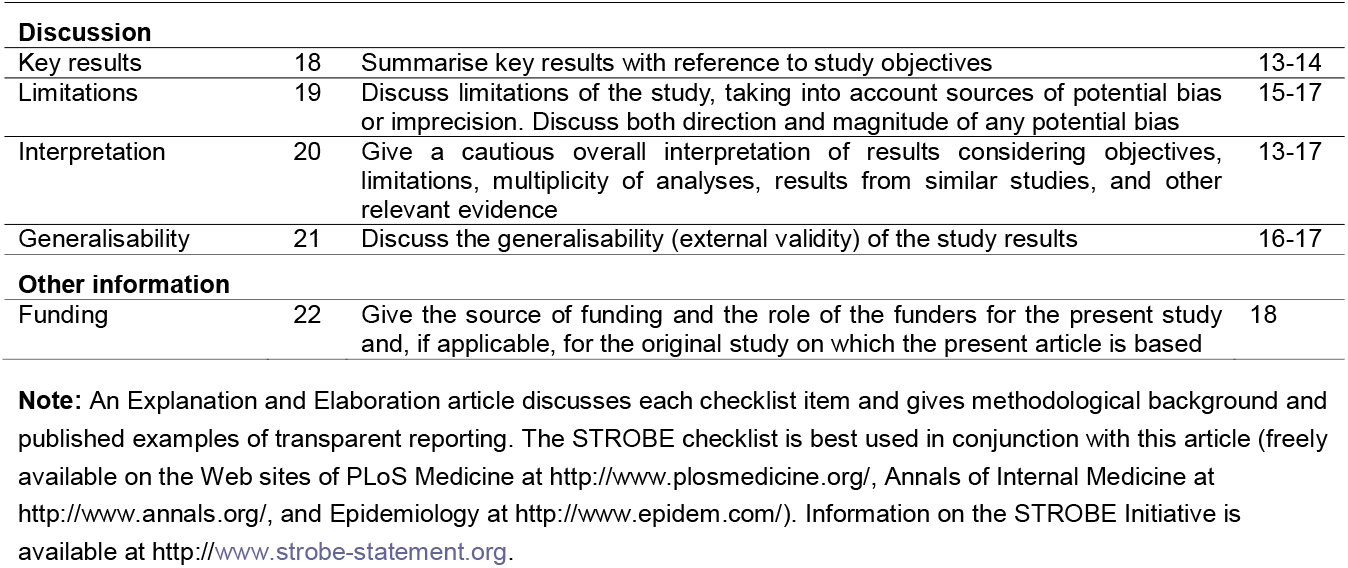

