## Supplementary for "Gastrointestinal Diseases, Genetic Risk, and Incident Dementia: A Prospective Cohort Study in 352,463 Middle-Aged Adults"

### **Supplementary Table 1.** ICD codes for identification of gastrointestinal diseases

| **Gastrointestinal** **disease** | **ICD-9** | **ICD-10** | **Reference** |
| --- | --- | --- | --- |
| **Esophagus** |  |  |  |
| Gastroesophageal reflux disease | 53011, 53081 | K21 | PMID: 17229223, 30548214 |
| Esophageal cancer | 150 | C15 | PMID: 31792601 |
| **Stomach and bowel** |  |  |  |
| Gastritis and duodenitis | 535 | K29 | PMID: 27194488 |
| Celiac disease | 5790 | K90.0 | PMID: 27501017 |
| Crohn’s disease | 555 | K50 | PMID: 27501017 |
| Ulcerative colitis | 556 | K51 | PMID: 27501017 |
| Intestinal diverticular disease | 562 | K57 | PMID: 34139333 |
| Irritable bowel syndrome | 5641 | K58 | PMID: 27501017, 31567167 |
| Peptic ulcer | 531-534 | K25-K28 | PMID: 32628718, 28503076, 32994689 |
| Gastric cancer | 151 | C16 | PMID: 20551458 |
| Small intestinal cancer | 152 | C17 | PMID: 31040384 |
| Colorectal cancer | 153, 1540, 1541 | C18-C20 | PMID: 22510213, 33675346 |
| **Pancreas** |  |  |  |
| Acute pancreatitis | 5770 | K85 | PMID: 34129395 |
| Chronic pancreatitis | 5771 | K86.0, K86.1 | PMID: 30039239 |
| Pancreatic cancer | 157 | C25 | PMID: 21346976 |
| **Gallbladder and biliary** |  |  |  |
| Cholangitis | 5761 | K83.0 | PMID: 27194488 |
| Cholecystitis | 575.0, 575.1 | K81 | PMID: 27194488 |
| Cholelithiasis | 574.0, 574.1, 574.2 | K80 | PMID: 27194488 |
| Gallbladder and biliary cancer | 156 | C23, C24 | PMID: 27516528, 31339558 |
| **Liver** |  |  |  |
| Non-alcoholic fatty liver disease | 5718, 5719 | K76.0, K75.8 | PMID: 26274335 |
| Cirrhosis | 5715, 5712, 4560, 4561, 4562, 5723, 5724, 5722, | K74.6, K70.3, I85, I98.2, I98.3, K76.6, K72.9, K76.7 | PMID: 35166399 |
| Liver cancers | 155 | C22 | PMID: 23182222, 23489585, 31339558 |
| **Appendix** |  |  |  |
| Appendicitis | 540-542 | K35-K37 |  |

ICD, international classification of disease

### **Supplementary Table 2.** Diagnostic codes (read codes) for identification of gastrointestinal diseases in primary care datasets

| **Gastrointestinal** **disease** | **Read codes version 2** | **Read codes version 3** |
| --- | --- | --- |
| **Esophagus** |  |  |
| Gastroesophageal reflux disease | J1011, J10y4 | J101., J1010, J1013, X76cn, J1011, J1012, J1020, X3003, X3004, X3009, XE0aL, Xa1q7, J1020, J10y4, Ua1kQ, X3003, X76cn, XE0aO, Xa7TZ, Xa7Ta, Xa7Tb |
| Esophageal cancer | B100., B10.., B101., B102., B103., B104., B105., B107., B106., B10y., B10z. | B10.., X300R, X78g3, XE1x5, XaB1b, XaFrD, B100., X78MH, B101., X78MM, B102., X78MR, B103., X78MW, XE1wz, B104., X78Mb, XE1x1, B105., X78Mg, XE1x3, XaOrV, B106., B10.., B10y., B10z., X300R, X78g3, XE1vQ, XE1x5, Xa0DF, Xa0DG, XaB1b, XaFrD, XaFrt |
| **Stomach and bowel** | |  |
| Gastritis and duodenitis | J15.., J1500, J150., Jyu12, J153., J1512, J1510, J151., J1511, J151z, J152., J154., J1540, J1541, J1542, J1543, J154z, J4300, J4z0., Jyu13, J1544, J155., J157., J156., J15z. | J15.., XaIN7, J150., X301N, X301O, X301R, X30Be, XaB3J, XaB3K, J150., Jyu12, X301N, X301P, X301R, J153., X301N, J151., J1512, X301N, X301R, J151., J1510, X301N, X301R, XE2bH, J151., J1511, J151z, X301N, J152., J154., J1541, J1542, J1543, J154z, J4300, J4z0., Jyu13, X301N, X301Q, X301R, X301S, X301T, X301U, X301V, X301W, X301X, X301Y, X301Z, X301a, XE0aT, XE2bH, J155., X301N, Xa356, XaIN7, J157., X3029, X302H, X302I, J15.., J156., J15z., X3029, Xa7yb |
| Celiac disease | J690., J6900, J6901, J690z | J690., J6900, J6901, J690z, X3034, X3035, X3037, X3039, X40Qn, XE0bK, XE0bL, XM1On |
| Crohn’s disease | J40.., J400., J4000, J4001, J4002, J4003, J4004, J4005, J400z, J401., J4010, J4011, J4012, J401z, J08z9, J402., Jyu40, J40z., N0311 | J5109, XE0ae, XE2QL, Xa0lh, Xa8Eh, J400., J4000, J4001, J4002, J4003, J4004, J400z, X302r, X302s, X302t, X302u, XaK6C, J401., J4010, J4011, J401z, X3050, XE0af, XaK6D, J08z9, J400., J4000, J4002, J4010, J4011, J402., Jyu40, X20Pq, X300J, X301b, X302r, X302t, J40z., J5109, XE2QL, Xa0lh, N0311, N0311, N0453, N0453, X701t, X701t, X7021, X7021, X702C, X702C, N0311, N0311, N0453, N0453, X701t, X701t, X7021, X7021, X702C, X702C |
| Ulcerative colitis | J4100, J411., J412., J413., J4103, J4102, J41y0, J438., J410., J41y., J41yz, J41z., Jyu41, J41.., J4101, J4104, J410z, N0310 | J410., X302z, X304H, XE0ae, XE0ag, Xa8Eh, XaK6E, J4100, X302s, X3030, XaB5z, XaYzX, J4103, X303e, J41y0, XaZ2j, J41y., J41yz, J41z., Jyu41, X302y, J41.., J410., J410z, X302y, XE0ag, XaK6E, N0310, N0310, N0454, N0454, X7021, X7021, X702C, X702C, N0310, N0310, N0454, N0454, X7021, X7021, X702C, X702C |
| Intestinal diverticular disease | J5120, J5121, J5122, J5123, J5124, J51.., J510., J511., J5100, J5101, J5102, J5103, J5104, J5110, J5111, J5112, J5113, J5114, J5125, J5126, J5127, J5105, J5106, J5107, J5115, J5116, J5117, J5128, J5108, J512., J512y, J512z, J513., J5109, J510y, J510z, J511y, J511z, J51z. | X302f, X3056, X3058, X305A, XE2sf, XE2sg, XaB8Z, J512., J5120, J5121, J5122, J5123, J5124, J57yB, X303Y, X303a, X304R, XM0B9, XM0v6, Xa1bJ, Xa1bM, Xa1yu, Xa1z1, Xa1z2, J5100, J5101, J5102, J5103, J5104, J5110, J5111, J5112, J5113, J5114, X303Y, XM0B8, XM0v6, Xa1bJ, Xa6p2, J512., J5125, J5126, J5127, J57yB, X304Q, X304R, X304T, XM0B9, Xa1bK, Xa1bL, Xa1d1, Xa9Fp, J5105, J5106, J5107, J5115, J5116, J5117, X304Q, X304T, XM0B8, Xa1bK, Xa1d1, Xa2ku, J512., J5128, J57yB, X303a, X304R, XM0B9, Xa1bJ, Xa1bK, Xa1bL, Xa1bM, Xa1yu, Xa1z1, Xa1z2, Xa2ku, Xa9Fp, J5108, X303Y, X304T, XM0B8, Xa1bJ, Xa1bK, J512., J512y, J512z, J51z., J55.., J57yB, X304R, XM0B9, J55.., J55.., J510y, J510z, J511y, J511z, X304P, XE2sf, XE2sg, XM0B8, XaB8Z |
| Irritable bowel syndrome | J5210, J5212, J521., J5211 | X302f, J5210, X3060, XE0as, XM0z0, XabiQ, J4..., J521., X305z, X3061, X76dS, XE0as, XM0z0, XabiP |
| Peptic ulcer | J11.., J112., J113., J1101, J1102, J1103, J110., J1100, J1104, J110y, J110z, J1111, J11y1, J1112, J11y2, J1113, J11y3, J111., J1110, J1114, J111y, J111z, J112z, J113z, J11y., J11y0, J11y4, J11yy, J11yz, J11z., J12.., J125., J126., J1201, J122., J1202, J1203, J120., J1200, J1204, J120y, J120z, J1211, J12y1, J12y., J1212, J12y2, J1213, J12y3, J121., J1210, J1214, J121y, J121z, J123., J124., J125z, J126z, J12y0, J12y4, J12yy, J12yz, J12z., J1301, J1302, J1303, J130., J1300, J1304, J130y, J130z, J1311, J13y1, J1312, J13y2, J1313, J13y3, J131., J1310, J1314, J131y, J131z, J13.., J13y., J13y0, J13y4, J13yy, J13yz, J13z., J14.., J1401, J1402, J1403, J140., J1400, J1404, J140y, J140z, J1411, J14y1, J1412, J14y2, J1413, J14y3, J141., J1410, J1414, J141y, J141z, J14y., J14y0, J14y4, J14yy, J14yz, J14z. | J1020, J11.., J11y., J11y0, J11y4, J11yz, X301G, X301L, X301M, X302X, X30Be, XE0aP, XE0aQ, XM0BZ, Xa6ot, Xa7nW, Xa84h, XaELE, XaLWq, XaLWr, XaMO5, XaMO6, J110., J1101, J1104, J110y, J110z, J130., J1301, J1304, J130y, J680., X301E, X301F, X301O, X301u, X30Bg, XE0bI, Xa1dj, XaB1x, XaB3J, XaB3K, XaBel, J110., J1102, J1104, J110y, J110z, J11y2, J130., J1302, J1304, J130y, X301E, X301F, X301o, J110., J1101, J1102, J1103, J1104, J110y, J110z, J11y2, J11y3, J130., J1301, J1302, J1303, J1304, J130y, X301E, X301F, X301O, X301o, X30Bg, Xa1dj, XaB1x, XaB3J, XaB3K, XaBel, J110., J1100, J1104, J110y, J110z, J130., J1300, J1304, J130y, X301E, X301F, X301H, X301I, J111., J1111, J1114, J111y, J111z, J11y1, J680., X301J, X301K, X301O, X30Bg, X76fI, XE0bI, Xa1dj, XaB1x, XaB3J, XaB3K, XaBel, J111., J1112, J1114, J111y, J111z, J11y2, X301J, X301K, X301n, X301o, J111., J1112, J1113, J1114, J111y, J111z, J11y1, J11y2, J11y3, X301J, X301K, X301O, X301n, X301o, X30Bg, Xa1dj, XaB3J, XaB3K, XaBel, J111., J1110, J1114, J111y, J111z, X301J, X301K, J1020, J11.., J11y., J11y0, J11y4, J11yy, J11yz, J11z., X301G, X301L, X301M, X302X, XE0aP, XE0aQ, Xa6ot, Xa7nW, Xa84h, XaELE, XaLWq, XaLWr, XaMO5, XaMO6, J12.., J124., J12y., J12y0, J12y4, J12yy, J12yz, J12z., X3029, X302B, X302G, X302H, X302b, X302c, X302w, X302z, X304H, X30Be, XM0BZ, XaLWs, XaLWt, XaMO7, XaMO8, J120., J1201, J1204, J120y, J120z, J12y1, J130., J1301, J1304, J130y, J680., X302A, X302C, X302D, X30Bh, XE0bI, Xa1dj, XaB3J, XaB3K, J120., J1202, J1204, J120y, J120z, J12y2, J130., J1302, J1304, J130y, X302A, X302C, X302D, X302Q, X303a, J120., J1201, J1202, J1203, J1204, J120y, J120z, J12y1, J12y2, J12y3, J130., J1301, J1302, J1303, J1304, J130y, X302A, X302C, X302D, X302Q, X303a, X30Bh, Xa1dj, XaB3J, XaB3K, J120., J1200, J1204, J120y, J120z, J130., J1300, J1304, J130y, X302A, X302C, X302D, J121., J1211, J1214, J121y, J121z, J12y1, J680., X302F, X30Bh, X76fI, XE0bI, Xa1dj, XaB3J, XaB3K, J121., J1212, J1214, J121y, J121z, J12y2, X302F, X302Q, X303a, J121., J1211, J1212, J1213, J1214, J121y, J121z, J12y1, J12y2, J12y3, X302F, X302Q, X303a, X30Bh, Xa1dj, XaB3J, XaB3K, J121., J1210, J1214, J121y, J121z, X302F, J12.., J124., J12y., J12y0, J12y4, J12yy, J12yz, J12z., X302B, X302G, X302b, X302c, XaLWs, XaLWt, XaMO7, XaMO8, X302w, X302z, X304H, X30Be, XM0BZ, XaYMi, J130., J1301, J1304, J130y, J680., XE0bI, Xa1dj, XaB3J, XaB3K, XaBmb, J130., J1302, J1304, J130y, XM0sI, J130., J1301, J1302, J1303, J1304, J130y, XM0sI, Xa1dj, XaB3J, XaB3K, XaBmb, J130., J1300, J1304, J130y, J130z, J1311, J13y1, J680., X76fI, XE0bI, XM1VW, Xa1dj, XaB3J, XaB3K, XaBmb, J1312, J13y2, X301n, XM0sI, J1313, J13y3, X301n, XM0sI, Xa1dj, XaB3J, XaB3K, XaBmb, J131., J1310, J1314, J131y, J131z, J13.., J13y., J13y0, J13y4, J13yy, J13yz, J13z., XE0aR, XM0BZ, XaYMi, J14.., X302w, X302x, X302z, X304H, X30Be, X40Ys, XE0aS, XM0BZ, Xa1qA, Xa1qB, Xa1qC, J130., J1301, J1304, J130y, J1401, J680., XE0bI, Xa1dj, XaB3J, XaB3K, J130., J1302, J1304, J130y, J1402, Xa1yt, J130., J1301, J1302, J1303, J1304, J130y, J1403, Xa1dj, Xa1yt, XaB3J, XaB3K, J130., J1300, J1304, J130y, J140., J1400, J1404, J140y, J140z, J1411, J14y1, J680., X76fI, XE0bI, Xa1dj, XaB3J, XaB3K, J1412, J14y2, X301n, Xa1yt, J1413, J14y3, Xa1dj, Xa1yt, XaB3J, XaB3K, J141., J1410, J1414, J141y, J141z, J14.., J14y., J14y0, J14y4, J14yy, J14yz, J14z., X302x, X40Ys, XE0aS, Xa1qA, Xa1qB, Xa1qC |
| Gastric cancer | B110., B1100, B1101, B110z, B118., B11.., B113., B114., B119., B112., B111., B1110, B1111, B111z, B115., B116., B117., B11y0, B11y1, B11y., B11yz, B11z. | B115., B116., X78gA, XE1vR, XE1xJ, Xa0DP, Xa0DQ, XaFrE, B110., B1100, B1101, B110z, X78Mm, XE1x7, XaOqX, B113., X78Mr, XE1xB, XE1xF, XE1xH, B114., X78Mw, XE1xD, XE1xF, XE1xH, XaOrB, B112., X78N1, XE1x9, XE1xF, XE1xH, B111., B1110, B1111, B111z, X78N6, XE1xF, XE1xH, B115., X78NB, XE1vS, XE1xF, B116., X78NG, XE1vT, XE1xH, B117., B11.., B11y., B11y0, B11y1, B11yz, B11z., X78gA, XE1vR, XE1xJ, Xa0DO, Xa0DP, Xa0DQ, XaBAo, XaFrE, XaFrs |
| Small intestinal cancer | B120., B12.., B121., B122., B123., B124., B12y., B12z. | X78gK, X78gL, B120., X78NL, B121., B122., XaA0C, B123., B124., B12.., B12y., B12z., X78gL, Xa0Df |
| Colorectal cancer | B134., B13.., B135., B136., B130., B131., B137., B132., B133., B138., B13y., B13z., PKyQ., B139., B140., B141. | B13.., X78gK, X78gN, X78gO, XE1xL, XE1xd, Xa0Dg, XaFrI, XaFsw, B134., X78gM, XE1vU, XaA0C, B135., B136., XE1xX, XaDc5, B130., XE1xN, XaDc8, B131., XE1xP, XaDc6, B137., X78e6, XE1xZ, XaDc9, B132., XE1xR, XaDc7, B133., XE1xT, Xa34H, Xa84V, B138., B13.., B13y., B13z., X78gN, X78gO, XE1vV, XE1xL, XE1xd, Xa0Dg, XaFrI, XaFro, XaFsw, B13.., B140., X78gK, X78gO, XE1vW, XE1xL, XE1xh, XaFrI, XaFrJ, B141., X78OK, XE1vW, XE1xf, Xa0Dj, XaFrJ, XaFrn |
| **Pancreas** |  |  |
| Acute pancreatitis | J670., J6706, J6705, J6707, J6708, J6709, J6701, J670y, J6700, J6702, J6703, J6704, J670z | J67.., J670., J6701, J6702, J6703, J6704, X308h, X308m, X3092, X3093, X3094, X3095, Xa0Rc, XaB1f, X308k, X308i, XaYaK, X308j, X308l, X308n, X308r, X308s, J670., J6700, J6701, J6702, J6703, J6704, X308h, X308m, X3092, XE0bH, Xa0Rc, XaB1f |
| Chronic pancreatitis | J6710, J671., Jyu84, J6711 | J671., X308h, X308v, Xa7na, J671., Jyu84, X308h, X308u, X308w, X308x, X308y, X308z, Xa7na |
| Pancreatic cancer | B170., B17.., B171., B172., B173., B174., B176., B17y., B17yz, B175., B17y0, B17z. | B17.., X78Ph, X78gd, XE1y5, XaFrH, XaFrp, B170., XE1xz, XaB1g, B171., XE1y1, XaB1h, B172., XE1y3, XaB1i, B173., B174., X50GW, X77nX, X78Pf, X78Pg, X78cy, X78cz, X78d1, X78d2, X78d3, Xa98N, XaJgM, B17y., X309C, X78gc, B175., B17.., B17y0, B17yz, B17z., X309D, X78Ph, X78gd, XE1y5, XaFrH, XaFrp |
| **Gallbladder and biliary** | |  |
| Cholangitis | J661., J6610, J6611, J6612, J6613, J6614, J6615, J6616, J6617, J6618, J6619, J661y, J661z | J661., J6610, J6611, J6612, J6613, J6614, J6615, J6616, J6617, J6618, J6619, J661y, J661z, X306W, X3086, Xa1k1, XaWzz |
| Cholecystitis | J650., J6500, J6501, J6502, J6503, J6504, J650z, J6510, J651., J651y, Jyu81, J651z | J65.., X3088, X3089, J650., J6500, J6501, J6502, J6503, J6504, J650z, X308B, XE0bF, XE2wM, XaB3H, XaE01, J6510, J651., J651y, Jyu81, J651z, X3089 |
| Cholelithiasis | J64.., J640., J6400, J6401, J640z, J641., J6410, J6411, J641z, J642., J6420, J6421, J642z, J64z., J64zz, J646., J643., J6430, J6431, J643z, J644., J6440, J6441, J644z, J6422, J645., J6450, J6451, J6452, J645z, J64z0, J64z1, Jyu80 | J65.., J6502, J6503, J6504, J6522, J6523, J6611, J6612, J6616, J662., J66y4, X3088, XE0bD, XE0bF, XaAwd, 4G2.., J640., J6400, J6401, J640z, X3089, X308A, XaB3H, 4G2.., J641., J6410, J6411, J641z, X3089, X308A, XaFr2, 4G2.., 4G21., 4G22., J64.., J642., J6420, J6421, J642z, J64z., J655., J6551, J6553, X308A, X308I, X308u, XE0bD, XM0zT, XaB3I, XaDc4, J646., J661., X308J, X308K, XaAzY, XaE6v, J643., J6430, J6431, J643z, J644., J6440, J6441, J644z, J661., X3089, X308J, XaB3H, 1965., 4G2.., J645., J6450, J6451, J645z, J6550, J6552, J664., J6640, X308J, X308K, XE0bE, XM0zT, Xa6bl, XaAzY, XaDc4, XaE6v, J64z0, J64z1, Jyu80 |
| Gallbladder and biliary cancer | B160., B16.., B161., B1610, B1611, B1612, B161z, B1613, B162., B163., B16y., B16z. | B160., X78gY, XE1vY, XE1xt, XaFrF, XaFrr, B16.., X78gb, B161., B1610, B1611, B1612, X77nj, X78PC, X78ga, XE1xv, XE2vN, B1613, B162., X78PX, XaFr0, B151., B163., B16.., B161z, B16y., B16z., X78gb, XaDbr |
| **Liver** |  |  |
| Non-alcoholic fatty liver disease | J61y1, J61y7, J61y9, J61y8, Jyu72 | Jyu72, X306p, X306q, XaQIT, J61y1, J61y7, X307v, XM095 |
| Liver cirrhosis | J6151, J6152, J6153, J6154, J6155, J6156, J6158, J6159, J615A, J615B, J615D, J615E, J615F, J615G, J615H, J615y, J615z, Jyu71, G8522, J612., G8523, G850., G851., G858., G852., G8521, G852z, Gyu94, G8520, J623., J624., J623., Xa9Fz | J61.., J615., J6150, J6151, J6152, J6153, J6154, J6155, J6156, J6158, J6159, J615A, J615B, J615D, J615E, J615F, J615G, J615H, J615y, J615z, Jyu71, X307L, X307M, X307N, X307O, X307P, X307Q, X307R, X307S, X307T, X307U, X307V, XE0b5, XE0b6, XE0b8, XE0bA, XM095, XaBM6, XaBM6, J612., J6152, J6155, X307L, X307M, X307Q, X307R, XE0b4, XE0b6, XE0b8, XM095, XaC1d, XaC1d, 2482., J102., X205y, X2063, XE0XW, XaE6u, G850., J10y0, Xa7TU, G851., X2063, XaE6u, G852., G8521, G852z, G852z, Gyu94, X205y, X2063, X2063, XaBM6, XaBM6, XaC1d, XaC1d, XaE6u, G852., G8520, G852z, G852z, Gyu94, X205y, X2063, X2063, XaBM6, XaBM6, XaC1d, XaC1d, XaE6u, J623., Xa9Fz, J624., J622., X0051, X0058, X3076, X3077, X3078, X3079, X307A, XE0bB, Xa01C, Xa8De, Xa8Df |
| Liver cancers | B1503, B15.., B15z., B151., B1510, B1511, B1512, B1513, B1514, B151z, B1501, B1502, Byu10, Byu11, B150., B1500, B150z, B152. | B150., X78Oz, XE1xp, Xa97q, B1500, B1503, X77nk, XM1FE, XaFrG, B151., B1510, B1511, B1512, B1513, B1514, B151z, B16.., B161., X78ga, X78gb, XE1xr, Xa97r, B1501, XaFrG, X78P0, XaFrG, Byu10, XaFrG, Byu11, XaFrG, B15.., B150., B150z, B152., B15z., X78Oz, XE1xp, Xa97q, XaFrG, XaFrq |
| **Appendix** |  |  |
| Appendicitis | J20.., J203., J200., J201., J204., J202., J20z., J20z1, J22.., J220., J221., J222., J223., J22z., Jyu20, J21.. | J21.., XE0cD, XM1Op, Xa9C3, Xa9C4, XaB9A, J20.., J20z1, J55.., J57yB, X304K, X304L, X309r, XA08f, Xa1z1, Xa3fg, Xa9Fp, XaYaL, XaZSs, XaZSu, J20.., J200., J20z1, J55.., J57yB, X304K, X304L, X304N, X309r, XA08f, XE0aV, Xa1z1, Xa3fg, Xa9Fp, XaYaN, XaZSs, XaZSu, J20.., J202., J20z., J20z1, J2y.., X304K, X304L, XA08f, J21.., J22.., J220., J221., J222., J223., J22z., Jyu20, XE0cD, XM1Op, Xa9C3, Xa9C4, XaB9A, J21.., XE0cD, XM1Op, Xa9C3, Xa9C4, XaB9A |

### **Supplementary Table 3.** Diagnostic codes for dementia and its subtypes

| **Source** | **Dementia** | **Alzheimer's disease** | **Vascular dementia** |
| --- | --- | --- | --- |
| ICD-9 | 3310, 3311, 290, 2900, 2901, 2902, 2903, 2904, 2908, 2909, 3334, 294, 2940, 2941, 2948, 2949, 293, 2930, 2931, 2938, 2939, 2921, 2922 | 3310 | 2904 |
| ICD-10 | F01, F010, F011, F012, F013, F018, F019, F02, F020, F021, F022, F023, F024, F028, F03, F051, F106, G30, G300, G301, G308, G309, G31, G310, G311, G312, G318, G319, I673, A810, F00, F000, F001, F002, F009 | F00, G30 | F01, I673 |
| Read v2 ^a^ | A411., A4110, F11x7, Eu021, F21y2, G678., F1100, Eu000, F1101, Eu001, Fyu30, Eu002, F110., Eu00., Eu00z, F11., F111., Eu020, F112., F11x0, F1440, F10y., F10y0, F10y1, F10y2, F10yz, F118., F11y., F11y2, F11yz, Fyu31, Eu025, F116., F10., F10z., F11xz, F11z., Eu0., Eu01., Eu010, Eu011, Eu012, Eu013, Eu01y, Eu01z, Eu02., Eu02y, Eu02z, Eu041, Eu106 | F1100, Eu000, F1101, Eu001, Fyu30, Eu002, F110., Eu00., Eu00z | Eu01., Eu010, Eu011, Eu012, Eu013, Eu01y, Eu01z, F21y2, G678. |
| Read v3 ^a^ | F110., F110., X002x, X002x, XaIKB, XaIKB, F110., F110., X0030, X0030, XaIKC, XaIKC, Eu002, Eu002, F110., F110., Eu00., Eu00., Eu00z, Eu00z, F110., F110., X00Qz, E0040, E0041, E0042, E0043, X00Qz, XE1Xs, Xa0lH, X003R, Xa0lH, XaIRJ, X003T, X003V, Eu01y, E0040, E0041, E0042, E0043, Eu01z, XE1Xs, Eu020, Eu020, F111., F111., X0034, X0034, Xa0s2, Xa0s2, Eu021, Eu021, XaA1S, XaA1S, XabVp, XabVp, Eu022, Eu022, Eu023, Eu023, X003l, X003l, XaOfZ, XaOfZ, X003P, X003P, Eu02., Eu02y, X003A, X003A, X003Y, X003Y, X00Dx, X00Dx, X00R2, X00R2, Xa0s2, Xa0s2, XaE74, XaE74, XaKyY, XaKyY, Eu02., X002w, X00Qz, E...., E000., E001., E0010, E0012, E0013, E002., E0020, E0021, Eu02z, X002w, X00Qz, X00R0, X00R2, X00SO, XE1Xr, XE1Z6, XM09N, XM09O, Xa0sE, XaX51, E0011, E003., E0041, Eu041, X002w, X00R2, XM09N, XM09O, 1B1A0, E011., E011., E0111, E0112, E011z, E02y2, E031., Eu106, X00RH, XE1Xv, XE1YQ, F110., X002y, X002z, XaIKB, F110., F110., X002x, X002x, XaIKB, XaIKB, X0031, X0032, XaIKC, F110., F110., X0030, X0030, X00R2, X00R2, XaIKC, XaIKC, Fyu30, X0033, Eu002, Eu002, F110., F110., F110., Eu00., Eu00., Eu00z, Eu00z, F110., F110., Xa0s2, Xa0s3, F111., X0035, X0036, X003E, Eu020, Eu020, F111., F111., X0034, X0034, Xa0s2, Xa0s2, F112., F11x0, F1440, X0056, XM0qi, XM0z3, Xa7nD, F10y., F10y0, F10y1, F10yz, F11yz, Fyu31, X002U, X002V, X002W, X0037, X0039, X003F, X003G, X003W, X003X, X003m, X004B, X004E, X005L, X005M, X005N, X005O, X005P, Xa0sC, XaPws, X003A, X003A, Xa0s2, Xa0s2, XaE74, XaE74, XaKyY, XaKyY, F1..., F10.., F10z., F11.., F11xz, F11z., X005K, X77qx, XE15F, Xa0s2, Xa0s3, Xa1GB, F21y2, A411., F11x7, X003K, XaA1S, XabVp, Eu021, Eu021, XaA1S, XaA1S, XabVp, XabVp | F110., F110., X002x, X002x, XaIKB, XaIKB, F110., F110., X0030, X0030, XaIKC, XaIKC, Eu002, Eu002, F110., F110., Eu00., Eu00., Eu00z, Eu00z, F110., F110., X00Qz, F110., X002y, X002z, XaIKB, F110., F110., X002x, X002x, XaIKB, XaIKB, X0031, X0032, XaIKC, F110., F110., X0030, X0030, X00R2, X00R2, XaIKC, XaIKC, Fyu30, X0033, Eu002, Eu002, F110., F110., F110., Eu00., Eu00., Eu00z, Eu00z, F110., F110. | E0040, E0041, E0042, E0043, X00Qz, XE1Xs, Xa0lH, X003R, Xa0lH, XaIRJ, X003T, X003V, Eu01y, E0040, E0041, E0042, E0043, Eu01z, XE1Xs, F21y2 |

^a^ diagnostic code for primary care data in UK.

### **Supplementary Table 4.** Genetic variants included in polygenic risk score

| **SNP** | **Chromosome** | **Position** | **Gene** | **Effect allele** | **Non-effect allele** | **Beta** | **Standard error** | **P value** |
| --- | --- | --- | --- | --- | --- | --- | --- | --- |
| rs4844610 | 1 | 207802552 | CR1 | A | C | 0.157 | 0.016 | 3.60E-24 |
| rs876461 | 2 | 37515958 | PRKD3/NDUFAF7 | G | A | -0.081 | 0.019 | 1.43E-05 |
| rs6733839 | 2 | 127892810 | BIN1 | C | T | -0.181 | 0.013 | 2.05E-44 |
| rs10933431 | 2 | 233981912 | INPP5D | G | C | -0.094 | 0.016 | 3.42E-09 |
| rs4351014 | 4 | 11027619 | HS3ST1 | T | C | 0.068 | 0.016 | 1.96E-05 |
| rs9275152 | 6 | 32652196 | HLA-DRB1 | T | C | 0.151 | 0.023 | 2.76E-11 |
| rs143332484 | 6 | 41129207 | TREM2 | C | T | -0.495 | 0.066 | 1.55E-14 |
| rs75932628 | 6 | 41129252 | TREM2 | C | T | -0.699 | 0.100 | 2.95E-12 |
| rs9381040 | 6 | 41154650 | TREML2 | C | T | 0.067 | 0.013 | 6.22E-07 |
| rs9381564 | 6 | 47443806 | CD2AP | A | G | -0.087 | 0.014 | 1.29E-10 |
| rs1859788 | 7 | 99971834 | PILRA | A | G | -0.082 | 0.014 | 1.22E-09 |
| rs56402156 | 7 | 143103481 | EPHA1 | G | A | 0.100 | 0.016 | 1.46E-10 |
| rs73223431 | 8 | 27219987 | PTK2B | C | T | -0.096 | 0.013 | 6.30E-14 |
| rs9331896 | 8 | 27467686 | CLU | C | T | -0.132 | 0.013 | 4.63E-24 |
| rs34674752 | 8 | 145154222 | SHARPIN | G | A | -0.107 | 0.042 | 1.19E-02 |
| rs34173062 | 8 | 145158607 | SHARPIN | G | A | -0.090 | 0.043 | 3.58E-02 |
| rs7920721 | 10 | 11720308 | ECHDC3 | A | G | -0.076 | 0.013 | 2.31E-09 |
| rs3740688 | 11 | 47380340 | SPI1h | G | T | -0.089 | 0.012 | 5.46E-13 |
| rs1582763 | 11 | 60021948 | MS4A | G | A | 0.114 | 0.013 | 2.36E-19 |
| rs3851179 | 11 | 85868640 | PICALM | T | C | -0.130 | 0.013 | 6.03E-25 |
| rs11218343 | 11 | 121435587 | SORL1 | T | C | 0.220 | 0.032 | 2.88E-12 |
| rs17125924 | 14 | 53391680 | FERMT2 | A | G | -0.128 | 0.021 | 1.42E-09 |
| rs11623019 | 14 | 92936971 | RIN3/SLC2A4 | T | C | 0.080 | 0.015 | 1.13E-07 |
| rs593742 | 15 | 59045774 | ADAM10 | A | G | 0.071 | 0.013 | 1.25E-07 |
| rs117618017 | 15 | 63569902 | APH1B | C | T | -0.094 | 0.026 | 2.38E-04 |
| rs7185636 | 16 | 19808163 | IQCK | T | C | 0.085 | 0.016 | 8.39E-08 |
| rs4985556 | 16 | 70694000 | IL34 | C | A | -0.091 | 0.020 | 3.82E-06 |
| rs12444183 | 16 | 81773209 | PLG2 | A | G | -0.054 | 0.013 | 1.74E-05 |
| rs3935877 | 16 | 81900853 | PLG2 | T | C | 0.062 | 0.023 | 7.21E-03 |
| rs72824905 | 16 | 81942028 | PLCG2 | C | G | 0.270 | 0.102 | 7.92E-03 |
| rs72835061 | 17 | 4805437 | CHRNE | C | A | -0.083 | 0.021 | 7.24E-05 |
| rs75511804 | 17 | 5138304 | SCIMP | C | T | -0.081 | 0.019 | 1.07E-05 |
| rs2732703 | 17 | 44353222 | KANSL1 | T | G | 0.069 | 0.026 | 7.89E-03 |
| rs616338 | 17 | 47297297 | ABI3 | T | C | 0.362 | 0.057 | 4.56E-10 |
| rs4311 | 17 | 61560763 | ACE | T | C | -0.048 | 0.014 | 8.18E-04 |
| rs3752231 | 19 | 1043638 | ABCA7 | C | T | -0.105 | 0.015 | 5.57E-12 |
| rs12459419 | 19 | 51728477 | CD33 | C | T | 0.080 | 0.016 | 4.51E-07 |
| rs6024870 | 20 | 54997568 | CASS4 | G | A | 0.123 | 0.022 | 3.46E-08 |
| rs2154481 | 21 | 27473875 | APP | C | T | -0.052 | 0.012 | 2.10E-05 |

### **Supplementary Table 5**. Definitions of covariates

| **Covariate** | **Definition** |
| --- | --- |
| Age at recruitment | This is a derived variable based on date of birth and date of attending an initial assessment center. It refers to the age of the participants on the day of attending an Initial Assessment Centre, truncated to whole year. |
| Sex | The phenotype (women and men) was determined by data from the National Health Service and self-reported questionnaires. |
| Educational attainment | From self-reported questionnaires: “Which of the following qualifications do you have” We classified the variable into: College (College or University degree) and Below college (A levels/AS levels or equivalent, O levels/GCSEs or equivalent, CSEs or equivalent, NVQ or HND or HNC or equivalent, other professional qualifications e.g., nursing, teaching, and none of the above) |
| Townsend deprivation index (TDI) | The higher, the more socioeconomic deprivation. TDI was derived according to the unemployment rate, the percentage of overcrowded households, the percentage of people without cars, and the percentage of people without houses for each area in the UK. The baseline TDI was calculated immediately before participant joining UK Biobank based on the preceding national census output areas. Each participant was assigned a score corresponding to the output area in which their postcode is located. |
| Smoking status | From self-reported questionnaires: current/past smoking status of the participant. We classified the variable into: Never smoked (never) and Ever smoker (previous and current) |
| Alcohol consumption | From self-reported questionnaires: Participants self-reported the number of alcohol units (10 ml of pure ethanol) consumed, in “units per week” (for frequent drinkers) or “units per month” (for less frequent drinkers), across several beverage categories (red wine, white wine/champagne, beer/cider, spirits, fortified wine, and “other”). To calculate alcohol consumption as per guidelines, multiply the volume by the alcohol content in percent and divide by drink-equivalent; then convert to grams.  1 drink-equivalent described as containing 14g of pure alcohol.  125ml wine=0.85 drink-equivalents,  4% ABV pint beer = 1.28 drink-equivalents,  25ml spirits=0.57 drink-equivalents,  50ml fortified wine= 0.56 drink-equivalents,  None-to-moderate level of alcohol consumption was defined as 0-14 g/d for women and 0-28 g/d for men according to US dietary guidelines, above which is defined as the heavy level. |
| Physical activity | From self-reported questionnaires: We classified the variable into Adequate (≥ 150 minutes moderate activity per week or ≥ 75 minutes vigorous activity per week or equivalent combination or moderate physical activity at least 5 days a week or vigorous activity once a week) and Inadequate (below adequate level) recommended by the American Heart Association. |
| Body biomass index (BMI) | BMI value is calculated from height and weight measured during the initial assessment centre visit. |
| Healthy diet | From self-reported questionnaires: Lourida I et al developed a healthy diet and associated the diet with dementia risk in the UK Biobank. Food frequency questionnaires collected following items in either quantitatively or as frequency of intake (e.g., 2-4 times/week). For the variables recorded in the frequency, we assigned it with mean value to get quantitative estimate (e.g., 2-4 times/week-> 3 servings/week). The individual was defined as with a healthy diet if the person met at least 4 of the following 7 requirements for food groups:  1. Fruits: ≥ 3 servings/day (items in FFQ: dried fruit, fresh fruit)  2. Vegetables: ≥ 3 servings/day (items in FFQ: cooked vegetable, raw or salad vegetable)  3. Fish: ≥2 servings/week (items in FFQ: oily fish, non-oily fish)  4. Processed meats: ≤ 1 serving/week (items in FFQ: processed meat)  5. Unprocessed red meats: ≤ 1.5 servings/week (items in FFQ: unprocessed pork, beef, mutton)  6. Whole grains: ≥ 3servings/day (items in FFQ: cereal intake, whole-meal or wholegrain bread)  7. Refined grains: ≤1.5servings/day (items in FFQ: other bread intake) |
| History of hypertension | The variable was coded as binary: yes or no with data from self-reported information, electronic health-related record, drug use, and baseline blood pressure measurement. |
| History of stroke | The variable was coded as binary: yes or no with data from self-reported information and electronic health-related record. |
| Family history | The variable was coded as binary: yes or no with data from self-reported Alzheimer's disease/dementia of parents and siblings. |
| History of depression | The variable was coded as binary: yes or no with data from self-reported: “Over the last 2 weeks, how often have you been bothered by feeling down, depressed, or hopeless”. We classified the variable into yes (Several days, More than half the days, Nearly every day, Several days) and no (“Not at all”) |

### **Supplementary Table 6.** Interaction and stratification analysis by sex and educational attainment

| **Gastrointestinal** **disease** | **Female** | **Male** | **P-interaction** | **Below college** | **College** | **P-interaction** |
| --- | --- | --- | --- | --- | --- | --- |
|  | **HR (95% CI)** | **HR (95% CI)** |  | **HR (95% CI)** | **HR (95% CI)** |  |
| Gastroesophageal reflux disease | 1.30 (1.16, 1.47) * | 1.63 (1.46, 1.82) * | 0.122 | 1.46 (1.34, 1.60) * | 1.42 (1.17, 1.71) * | 0.995 |
| Gastritis and duodenitis | 1.58 (1.38, 1.81) * | 1.80 (1.58, 2.06) * | 0.796 | 1.66 (1.50, 1.84) * | 1.76 (1.40, 2.21) * | 0.995 |
| Celiac disease | 1.52 (0.91, 2.52) | 1.25 (0.84, 1.87) | 0.831 | 1.48 (1.05, 2.09) * | 0.89 (0.40, 1.99) | 0.627 |
| Crohn’s disease | 1.16 (0.66, 2.04) | 1.60 (1.00, 2.59) | 0.796 | 1.20 (0.78, 1.85) | 2.34 (1.17, 4.70) | 0.627 |
| Ulcerative colitis | 1.01 (0.67, 1.53) | 1.11 (0.73, 1.69) | 0.831 | 0.98 (0.70, 1.38) | 1.40 (0.77, 2.53) | 0.627 |
| Intestinal diverticular disease | 1.08 (0.91, 1.28) | 1.45 (1.25, 1.68) * | 0.122 | 1.34 (1.19, 1.52) * | 0.95 (0.71, 1.26) | 0.402 |
| Irritable bowel syndrome | 1.61 (1.32, 1.96) * | 1.63 (1.43, 1.86) * | 0.907 | 1.56 (1.37, 1.77) * | 1.83 (1.48, 2.26) * | 0.627 |
| Peptic ulcer | 1.41 (1.20, 1.66) * | 1.59 (1.29, 1.95) * | 0.831 | 1.47 (1.28, 1.69) * | 1.45 (1.05, 2.01) | 0.995 |
| Pancreatitis | 1.42 (0.82, 2.45) | 1.22 (0.70, 2.10) | 0.831 | 1.48 (1.00, 2.19) | 0.33 (0.05, 2.32) | 0.402 |
| Gallbladder disease | 1.00 (0.78, 1.28) | 1.19 (1.02, 1.38) | 0.796 | 1.14 (0.99, 1.31) | 1.09 (0.81, 1.48) | 0.995 |
| Non-alcoholic fatty liver disease | 1.88 (1.11, 3.18) * | 2.22 (1.26, 3.92) * | 0.831 | 2.00 (1.30, 3.07) * | 2.05 (0.85, 4.95) | 0.995 |
| Cirrhosis | 3.23 (2.10, 4.97) * | 1.45 (0.82, 2.55) | 0.122 | 2.21 (1.51, 3.22) * | 2.28 (1.02, 5.08) | 0.995 |
| Appendicitis | 0.80 (0.57, 1.14) | 0.92 (0.66, 1.29) | 0.831 | 0.79 (0.59, 1.05) | 1.08 (0.70, 1.66) | 0.627 |
| Overall gastrointestinal cancer | 1.04 (0.73, 1.48) | 1.16 (0.76, 1.77) | 0.831 | 1.12 (0.83, 1.51) | 0.95 (0.51, 1.77) | 0.995 |

CI, confidence interval; HR, hazard ratio.

Estimates were adjusted for age, sex, Townsend deprivation index, educational attainment, BMI, physical activity, diet, smoking status, alcohol consumption, baseline hypertension, baseline stroke, and polygenic risk score. * Representing a significant association after FDR correction for multiple comparison. *P* for interaction was two-sided FDR-adjusted.

### **Supplementary Table 7**. Risk of incident dementia according to polygenic risk score

|  |  | **Model 1 ^a^** |  | **Model 2 ^b^** |  |
| --- | --- | --- | --- | --- | --- |
|  | **Case/Person-years** | **HR (95% CI)** | **P value** | **HR (95% CI)** | **P value** |
| **Polygenic risk score categories** | | |  |  |  |
| Low (quintile 1) | 928/825,184 | Ref |  | Ref |  |
| Intermediate (quintile 2-4) | 3307/2,476,237 | 1.19 (1.11, 1.28) | <0.001 | 1.19 (1.11, 1.28) | <0.001 |
| High (quintile 5) | 1413/82,454 | 1.53 (1.41, 1.66) | <0.001 | 1.54 (1.41, 1.67) | <0.001 |
| *P* value for trend ^c^ | |  | <0.001 |  | <0.001 |
| **In quintiles** |  |  |  |  |  |
| 1 (lowest) | 928/825,184 | Ref |  | Ref |  |
| 2 | 1061/824,684 | 1.15 (1.05, 1.25) | 0.002 | 1.14 (1.05, 1.25) | 0.003 |
| 3 | 1064/826,251 | 1.15 (1.05, 1.25) | 0.002 | 1.15 (1.05, 1.26) | 0.002 |
| 4 | 1182/825,302 | 1.27 (1.17, 1.39) | <0.001 | 1.28 (1.17, 1.39) | <0.001 |
| 5 (highest) | 1413/824,542 | 1.53 (1.41, 1.66) | <0.001 | 1.54 (1.41, 1.67) | <0.001 |
| *P* value for trend ^c^ | |  | <0.001 |  | <0.001 |

^a^ adjusted for the age, sex.

^b^ adjusted for age, sex, Townsend deprivation index, educational attainment, BMI, physical activity, diet, smoking status, alcohol consumption, baseline hypertension, and baseline stroke

^c^ The trend test used the median value of each group instead of the original group.

### **Supplementary Table 8**. Results of the analysis using competing risk model

| **Gastrointestinal** **disease** | **HR (95% CI)** | ***P*** | **Q value** |
| --- | --- | --- | --- |
| Gastroesophageal reflux disease | 1.44 (1.33, 1.55) | <0.001 | <0.001 |
| Gastritis and duodenitis | 1.63 (1.49, 1.78) | <0.001 | <0.001 |
| Celiac disease | 1.36 (1.00, 1.83) | 0.048 | 0.084 |
| Crohn’s disease | 1.30 (0.92, 1.85) | 0.140 | 0.196 |
| Ulcerative colitis | 1.03 (0.78, 1.36) | 0.860 | 0.860 |
| Intestinal diverticular disease | 1.26 (1.13, 1.40) | <0.001 | <0.001 |
| Irritable bowel syndrome | 1.59 (1.43, 1.76) | <0.001 | <0.001 |
| Peptic ulcer | 1.41 (1.25, 1.60) | <0.001 | <0.001 |
| Pancreatitis | 1.21 (0.84, 1.75) | 0.320 | 0.407 |
| Gallbladder disease | 1.13 (0.99, 1.28) | 0.065 | 0.101 |
| Non-alcoholic fatty liver disease | 1.70 (1.26, 2.50) | 0.007 | 0.014 |
| Cirrhosis | 1.79 (1.27, 2.52) | <0.001 | <0.001 |
| Appendicitis | 0.93 (0.75, 1.16) | 0.540 | 0.630 |
| Overall gastrointestinal cancer | 1.07 (0.84, 1.37) | 0.600 | 0.646 |

CI, confidence interval; HR, hazard ratio.

Associations were derived from model 2 adjusted for age, sex, Townsend deprivation index, educational attainment, BMI, physical activity, diet, smoking status, alcohol consumption, baseline hypertension, baseline stroke, and polygenic risk score.

### **Supplementary Table 9.** Sensitivity analyses for the associations between gastrointestinal diseases and risk of dementia

| **Gastrointestinal diseases** | **Excluding incident dementia in the first 3 year** | | | **Further adjusted for depression** | | | **Further adjusted for family history of dementia** | | | **Further adjusted for regular use of proton pump inhibitors** | | |
| --- | --- | --- | --- | --- | --- | --- | --- | --- | --- | --- | --- | --- |
|  | **HR (95% CI)** | ***P*** | **Q value** | **HR (95% CI)** | ***P*** | **Q value** | **HR (95% CI)** | ***P*** | **Q value** | **HR (95% CI)** | ***P*** | **Q value** |
| Gastroesophageal reflux disease | 1.37 (1.25, 1.49) | <0.001 | <0.001 | 1.41 (1.30, 1.53) | <0.001 | <0.001 | 1.46 (1.34, 1.58) | <0.001 | <0.001 | 1.38 (1.25, 1.51) | <0.001 | <0.001 |
| Gastritis and duodenitis | 1.57 (1.41, 1.74) | <0.001 | <0.001 | 1.62 (1.48, 1.79) | <0.001 | <0.001 | 1.68 (1.53, 1.85) | <0.001 | <0.001 | 1.62 (1.47, 1.79) | <0.001 | <0.001 |
| Celiac disease | 1.24 (0.87, 1.76) | 0.241 | 0.281 | 1.33 (0.97, 1.83) | 0.073 | 0.127 | 1.35 (0.98, 1.84) | 0.065 | 0.101 | 1.35 (0.98, 1.84) | 0.065 | 0.114 |
| Crohn’s disease | 1.45 (0.98, 2.13) | 0.061 | 0.106 | 1.35 (0.93, 1.94) | 0.110 | 0.154 | 1.39 (0.96, 2.00) | 0.077 | 0.108 | 1.33 (0.93, 1.92) | 0.122 | 0.190 |
| Ulcerative colitis | 1.21 (0.90, 1.62) | 0.210 | 0.267 | 1.03 (0.77, 1.38) | 0.836 | 0.836 | 1.06 (0.79, 1.42) | 0.706 | 0.706 | 1.04 (0.78, 1.40) | 0.775 | 0.775 |
| Intestinal diverticular disease | 1.25 (1.11, 1.40) | <0.001 | 0.001 | 1.23 (1.10, 1.38) | <0.001 | 0.001 | 1.26 (1.13, 1.41) | <0.001 | <0.001 | 1.28 (1.14, 1.43) | <0.001 | <0.001 |
| Irritable bowel syndrome | 1.47 (1.30, 1.66) | <0.001 | <0.001 | 1.55 (1.39, 1.72) | <0.001 | <0.001 | 1.62 (1.45, 1.81) | <0.001 | <0.001 | 1.56 (1.40, 1.74) | <0.001 | <0.001 |
| Peptic ulcer | 1.41 (1.22, 1.61) | <0.001 | <0.001 | 1.43 (1.26, 1.63) | <0.001 | <0.001 | 1.48 (1.30, 1.68) | <0.001 | <0.001 | 1.32 (1.15, 1.50) | <0.001 | <0.001 |
| Pancreatitis | 1.32 (0.87, 1.98) | 0.188 | 0.264 | 1.28 (0.87, 1.88) | 0.216 | 0.253 | 1.30 (0.89, 1.92) | 0.178 | 0.210 | 1.26 (0.86, 1.86) | 0.235 | 0.300 |
| Gallbladder disease | 1.06 (0.92, 1.21) | 0.449 | 0.449 | 1.11 (0.98, 1.26) | 0.103 | 0.154 | 1.13 (0.99, 1.29) | 0.060 | 0.101 | 1.10 (0.96, 1.25) | 0.164 | 0.230 |
| Non-alcoholic fatty liver disease | 1.94 (1.27, 2.94) | 0.002 | 0.004 | 1.97 (1.34, 2.90) | 0.001 | 0.001 | 2.03 (1.38, 2.99) | <0.001 | 0.001 | 1.95 (1.33, 2.87) | 0.001 | 0.001 |
| Cirrhosis | 1.93 (1.30, 2.86) | 0.001 | 0.002 | 2.19 (1.55, 3.08) | <0.001 | <0.001 | 2.22 (1.57, 3.12) | <0.001 | <0.001 | 2.28 (1.62, 3.22) | <0.001 | <0.001 |
| Appendicitis | 0.82 (0.63, 1.08) | 0.156 | 0.242 | 0.86 (0.68, 1.09) | 0.217 | 0.253 | 0.85 (0.67, 1.08) | 0.180 | 0.210 | 1.10 (0.86, 1.40) | 0.441 | 0.475 |
| Overall gastrointestinal cancer | 1.15 (0.87, 1.52) | 0.329 | 0.354 | 1.08 (0.82, 1.42) | 0.573 | 0.618 | 1.09 (0.83, 1.43) | 0.537 | 0.578 | 1.13 (0.86, 1.48) | 0.373 | 0.435 |

CI, confidence interval; HR, hazard ratio. Estimates were adjusted for age, sex, Townsend deprivation index, educational attainment, BMI, physical activity, diet, smoking status, alcohol consumption, baseline hypertension, baseline stroke, and polygenic risk score.

### **Supplementary Table 10.** Associations between gastrointestinal diseases and incident dementia in the analysis including individuals who developed subsequent gastrointestinal disease after baseline

|  | **Case/person-year** | **HR (95% CI)** | ***P*** | **Q value** |
| --- | --- | --- | --- | --- |
| **Gastroesophageal reflux disease** | | |  |  |
| Without | 7175/4,733,923 | Ref |  |  |
| With | 2446/889,169 | 1.46 (1.40, 1.53) | <0.001 | <0.001 |
| **Gastritis and duodenitis** | | |  |  |
| Without | 7501/4,941,041 | Ref |  |  |
| With | 2120/682,051 | 1.59 (1.51, 1.67) | <0.001 | <0.001 |
| **Celiac disease** | |  |  |  |
| Without | 9511/5,576,533 | Ref |  |  |
| With | 110/46,560 | 1.30 (1.07, 1.56) | 0.007 | 0.007 |
| **Crohn’s disease** | |  |  |  |
| Without | 9525/5,584,496 | Ref |  |  |
| With | 96/38,596 | 1.35 (1.10, 1.65) | 0.004 | 0.004 |
| **Ulcerative colitis** | |  |  |  |
| Without | 9442/5,551,227 | Ref |  |  |
| With | 179/71,865 | 1.30 (1.12, 1.51) | 0.001 | 0.001 |
| **Intestinal diverticular disease** | | |  |  |
| Without | 7569/4,884,578 | Ref |  |  |
| With | 2052/738,515 | 1.26 (1.20, 1.32) | <0.001 | <0.001 |
| **Irritable bowel syndrome** | | |  |  |
| Without | 8708/5,248,549 | Ref |  |  |
| With | 913/374,543 | 1.55 (1.45, 1.66) | <0.001 | <0.001 |
| **Peptic ulcer** | |  |  |  |
| Without | 8770/5,382,979 | Ref |  |  |
| With | 851/240,113 | 1.55 (1.44, 1.66) | <0.001 | <0.001 |
| **Pancreatitis** | |  |  |  |
| Without | 9429/5,572,728 | Ref |  |  |
| With | 192/50,364 | 1.73 (1.50, 2.00) | <0.001 | <0.001 |
| **Gallbladder disease** | |  |  |  |
| Without | 8480/5,216,357 | Ref |  |  |
| With | 1141/406,735 | 1.39 (1.31, 1.48) | <0.001 | <0.001 |
| **Non-alcoholic fatty liver disease** | | |  |  |
| Without | 9208/5,526,650 | Ref |  |  |
| With | 413/96,442 | 2.29 (2.07, 2.52) | <0.001 | <0.001 |
| **Cirrhosis** |  |  |  |  |
| Without | 9307/5,574,940 | Ref |  |  |
| With | 314/48,152 | 3.22 (2.88, 3.61) | <0.001 | <0.001 |
| **Appendicitis** | |  |  |  |
| Without | 9436/5,515,412 | Ref |  |  |
| With | 185/107,680 | 1.02 (0.88, 1.17) | 0.834 | 0.834 |
| **Overall gastrointestinal cancer** | | |  |  |
| Without | 9200/5,475,118 | Ref |  |  |
| With | 421/147,975 | 1.20 (1.09, 1.32) | <0.001 | <0.001 |

CI, confidence interval; HR, hazard ratio.

Estimates were adjusted for age, sex, Townsend deprivation index, educational attainment, BMI, physical activity, diet, smoking status, alcohol consumption, baseline hypertension, baseline stroke, and polygenic risk score.

### **Supplementary Table 11.** Associations between gastrointestinal diseases and incident dementia in the analysis excluding individuals with Parkinson’s disease (n=2265)

| **Gastrointestinal diseases** | **HR (95% CI)** | ***P*** | **Q value** |
| --- | --- | --- | --- |
| Gastroesophageal reflux disease | 1.52 (1.39, 1.65) | <0.001 | <0.001 |
| Gastritis and duodenitis | 1.74 (1.58, 1.93) | <0.001 | <0.001 |
| Celiac disease | 1.28 (0.91, 1.81) | 0.154 | 0.195 |
| Crohn’s disease | 1.36 (0.92, 2.01) | 0.128 | 0.180 |
| Ulcerative colitis | 1.05 (0.76, 1.43) | 0.779 | 0.779 |
| Intestinal diverticular disease | 1.31 (1.16, 1.47) | <0.001 | <0.001 |
| Irritable bowel syndrome | 1.70 (1.52, 1.90) | <0.001 | <0.001 |
| Peptic ulcer | 1.51 (1.32, 1.73) | <0.001 | <0.001 |
| Pancreatitis | 1.38 (0.93, 2.07) | 0.112 | 0.175 |
| Gallbladder disease | 1.15 (1.01, 1.31) | 0.042 | 0.073 |
| Non-alcoholic fatty liver disease | 2.29 (1.56, 3.36) | <0.001 | <0.001 |
| Cirrhosis | 2.39 (1.69, 3.39) | <0.001 | <0.001 |
| Appendicitis | 0.91 (0.71, 1.17) | 0.467 | 0.503 |
| Overall gastrointestinal cancer | 1.14 (0.85, 1.51) | 0.380 | 0.443 |

CI, confidence interval; HR, hazard ratio.

Estimates were adjusted for age, sex, Townsend deprivation index, educational attainment, BMI, physical activity, diet, smoking status, alcohol consumption, baseline hypertension, baseline stroke, and polygenic risk score.

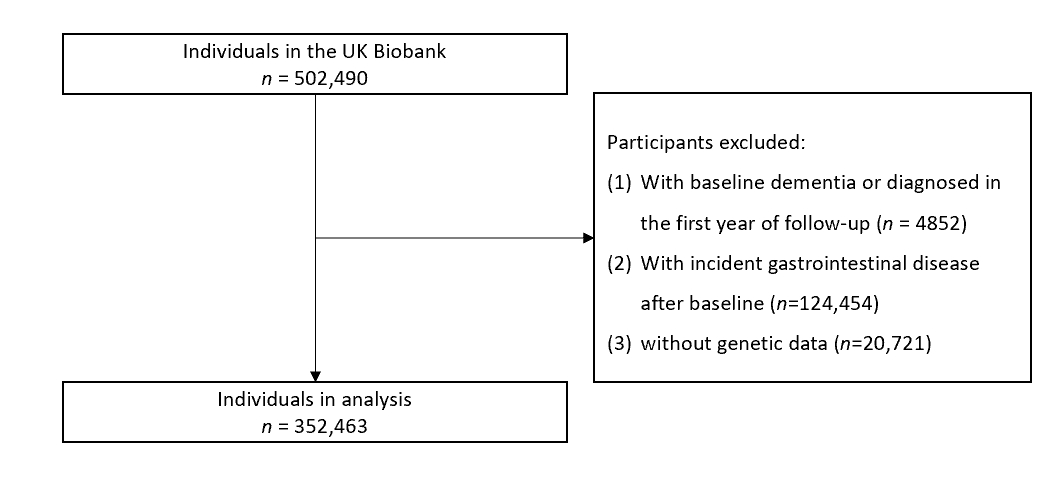

### **Supplementary Figure 1**. Flowchart of inclusion of study participants

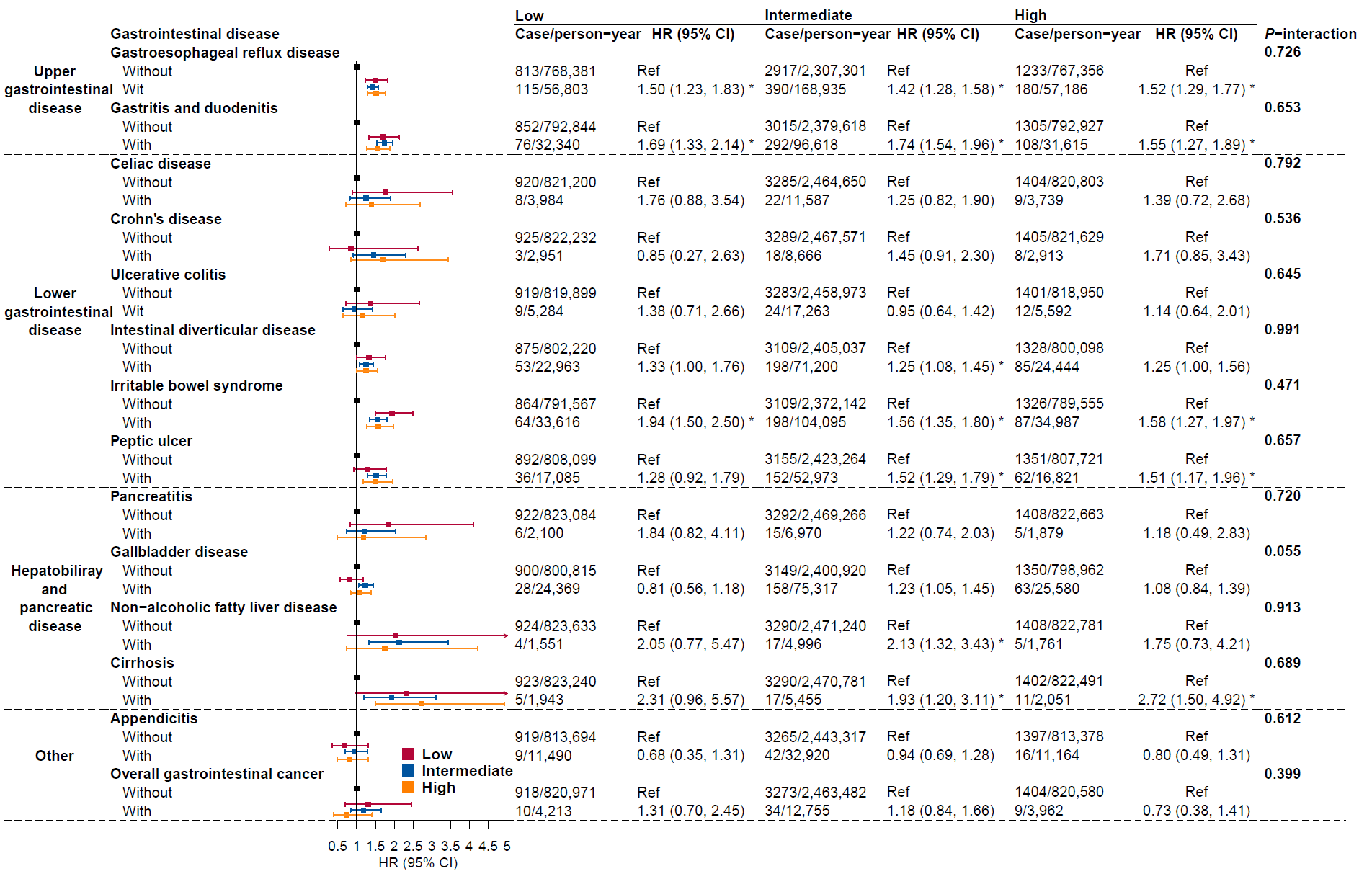

### **Supplementary Figure 2**. Associations between baseline gastrointestinal diseases and risk of incident dementia stratified by polygenic risk score categories. CI, confidence interval; HR, hazard ratio. Estimates were adjusted for sex, Townsend deprivation index, educational attainment, BMI, physical activity, diet, smoking status, alcohol consumption, baseline hypertension, baseline stroke. * Representing a significant association after FDR correction for multiple comparison. P for interaction was two-sided FDR-adjusted.

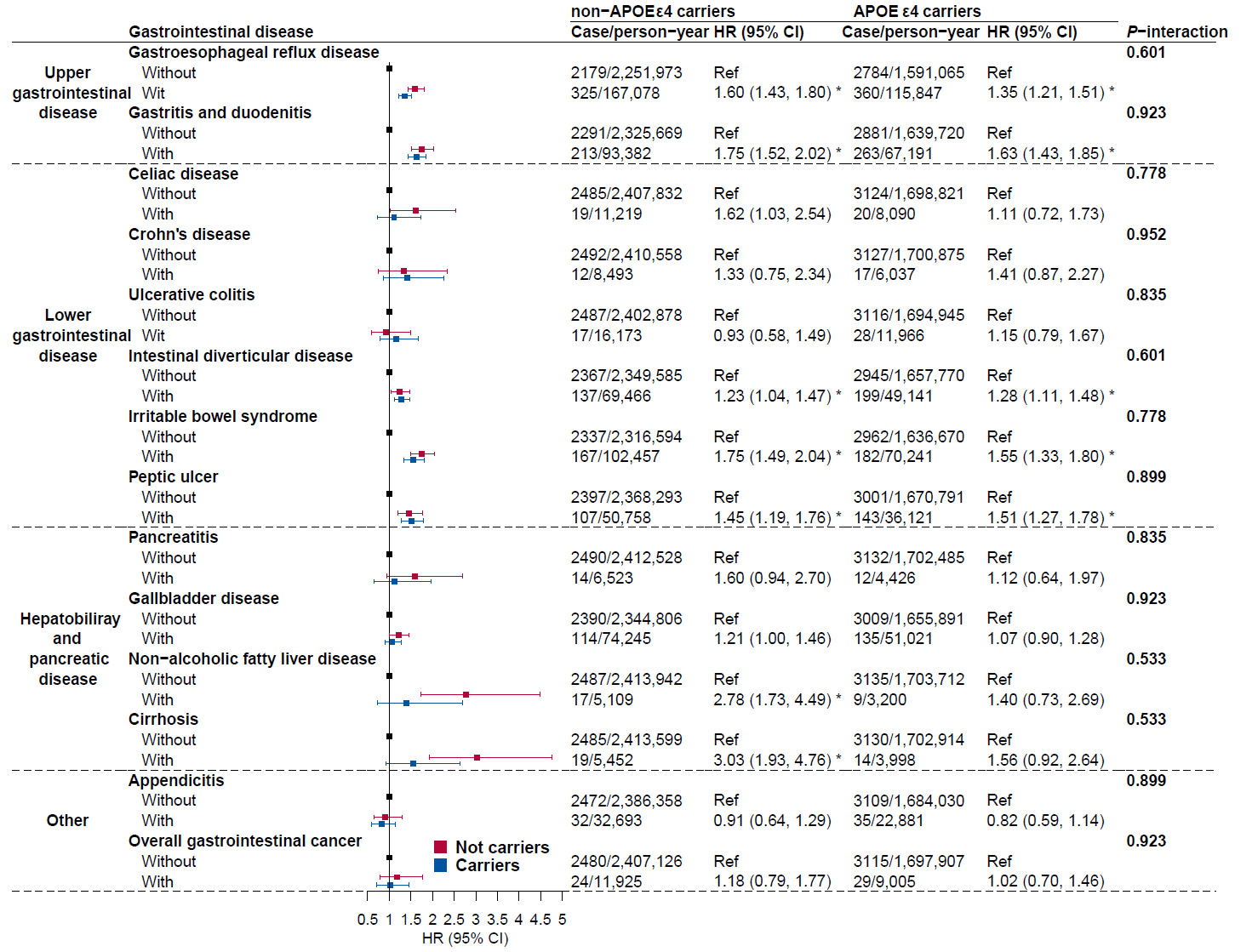

### **Supplementary Figure 3**. **Associations between baseline gastrointestinal diseases and risk of incident dementia stratified by *APOE* ε4 status.** CI, confidence interval; HR, hazard ratio. Estimates were adjusted for age, sex, Townsend deprivation index, educational attainment, BMI, physical activity, diet, smoking status, alcohol consumption, baseline hypertension, baseline stroke. * Representing a significant association after FDR correction for multiple comparison. *P* for interaction was two-sided FDR-adjusted.
